# Examining the independent roles of cannabis use and tobacco use in depression risk: a multivariable Mendelian randomisation study

**DOI:** 10.1101/2024.07.17.24310564

**Authors:** Chloe Burke, Tom P Freeman, Hannah Sallis, Robyn E Wootton, Gemma MJ Taylor

## Abstract

**Background:** Cannabis and tobacco use are consistently associated with major depressive disorder (MDD) in conventional observational studies. However, these substances are often co-used, and the independent causal role of cannabis use in risk of MDD remains unclear.

**Methods:** Univariable and multivariable MR (MVMR) were used to explore the total and independent causal effects of genetic liability to tobacco use and cannabis use on MDD. Our primary estimator was the inverse-variance weighted (IVW) method, with other methods as sensitivity analyses. For the exposures, we used genome-wide association study (GWAS) summary statistics among European ancestry individuals for several tobacco use (i.e., smoking initiation, smoking continuation, smoking heaviness) and cannabis use (i.e., cannabis initiation, cannabis use disorder [CUD]) phenotypes. For the outcome, a GWAS of MDD was conducted using individual-level data from UK Biobank.

**Results:** Univariable MR indicated a causal effect of smoking initiation on MDD (odds ratio [OR]^IVW^ = 1.34, 95% confidence interval [CI] = 1.27 – 1.42), with consistent but weaker evidence for smoking continuation (OR^IVW^ = 1.13, 95% CI = 0.93 – 1.37) and smoking heaviness (OR^IVW^ = 1.15, 95% CI = 0.99 – 1.33). There was no clear evidence for a causal effect of cannabis initiation on MDD (OR^IVW^ = 1.00, 95% CI = 0.91– 1.11). Univariable MR indicated some evidence for a causal effect of CUD on MDD (OR^IVW^ = 1.14, 95% CI = 1.04 – 1.25), which attenuated to the null when adjusting for liability to smoking initiation (OR^MVMR-IVW^ = 1.03, 95% CI = 0.97 – 1.08).

**Conclusions:** This study provides limited evidence for an independent causal effect of cannabis use on MDD, and stronger evidence for an independent causal effect of tobacco use on MDD. Analyses were limited by low power, and future research should triangulate these findings with results from high-quality observational studies.

## BACKGROUND

Major Depressive Disorder (MDD) is a highly prevalent psychiatric disorder (e.g., lifetime prevalence ∼11%) [1]. In terms of global disease burden, MDD was estimated as a leading cause of years lived with disability (YLDs) [2]. However, knowledge of actionable preventative strategies that could mitigate depression risk remains limited [3].

Tobacco smoking and cannabis use are both prospectively associated with increased risk of depression in conventional observational studies [4–10]. These substances are commonly co-used, a behaviour comprising ‘concurrent use’ (i.e., use of both products in a pre-defined time period, including ‘sequential’ use) and co-administration (i.e., used simultaneously via the same delivery method) [11], the latter of which is more common in European countries [12]. Despite their consistent association with depression, there are multiple limitations to inferring causality from conventional observational studies (e.g., reverse causation, confounding bias) [13,14]. Particularly confounding from other substance use, making it crucially important to disentangle the independent causal effect of cannabis use from tobacco use, and vice versa.

Mendelian randomisation (MR) is an epidemiological approach which employs genetic variants as proxies for levels of an exposure in an instrumental variable analysis [15]. Genetic variants which alter the average lifetime levels of an exposure are randomised at conception and inherited independently of conventional confounding factors [16]. As such, MR should be more robust to key sources of bias than conventional observational studies [16,17]. MR requires three key assumptions to be met of the genetic instruments to yield a valid causal estimate; (i) it is robustly associated with the exposure (i.e., ‘relevance’), (ii) it does not share a common cause with the outcome (i.e., ‘exchangeability’), and (iii) it affects the outcome only through the exposure (i.e., ‘exclusion restriction’) [16].

Previous MR studies have yielded mixed evidence regarding the effect of these substances on depression [18]. More recent studies, using improved genetic instruments (e.g., lifetime smoking index) and larger samples, suggest a causal role of tobacco smoking on depression [18,19]. In contrast, there is limited evidence for causal effects of cannabis use on depression [20]. This could suggest that prospective observational associations are biased by unmeasured confounding, including tobacco use [21]. However, this may also relate to the lack of availability of cannabis use instruments beyond initiation of use [18,22]. A review of MR studies examining substance use and mental health suggested several key improvements for future MR studies in this area, including: (i) additional sensitivity analyses; (ii) use of phenotypes which characterise heaviness of use; and (iii) use of multivariable MR [18].

Multivariable MR (MVMR) is an extension to MR that includes multiple exposures to estimate the effect of one exposure independent of other, genetically correlated, exposures [23,24]. MVMR is therefore a valuable tool to explore highly correlated phenotypes, such as tobacco smoking and cannabis use which could result in horizonal pleiotropy (i.e., genetic variants influencing the outcome independently of the target exposure) [25], if not accounted for in the same model [24]. This method has been previously applied to disentangle the role of cannabis and tobacco use in schizophrenia [26] and suicide attempt [27], but is yet to be applied to examine the independent effects of these substances on depression.

In this study, we applied univariable and multivariable MR to estimate the independent effect of cannabis use (versus tobacco smoking) on major depressive disorder (MDD).

## METHODS

### Study overview

This study employed univariable and multivariable MR with summary-level data to examine the total and independent causal effects of tobacco smoking and cannabis use on MDD. A statistical analysis plan was pre-registered (https://osf.io/bg9vk/), and deviations have been reported in Supplementary Note **S1**.

### Ethics statement

UK Biobank received ethics approval from the North West Multi-Centre Research Ethics Committee as a Research Tissue Bank approval (REC; 11/NW/0382). Approval to use these data was sought and approved by UK Biobank (Project ID: 9142).

### Data Sources

Summary-level genetic data obtained from genome-wide association studies (GWAS) were used to identify relevant instrumental variables (IVs). For the exposures, we used SNPs associated with smoking initiation [28], smoking continuation [28], smoking heaviness (measured via cigarettes per day) [28], cannabis initiation [29] and cannabis use disorder (CUD) [30]. Information regarding the genotyping, imputation and quality control of each sample is reported in the original studies [28–30]. The GWAS of smoking continuation and heaviness were performed amongst ever smokers [28]. Therefore, we required the summary-level statistics for the outcome to be stratified by smoking status in order to be comparable with the underlying population of the exposure GWAS data [15,31]. We were not aware of any existing available GWAS summary-level data for MDD appropriately stratified by smoking status. As such, we used UK Biobank data to conduct a GWAS of major depressive disorder (MDD) stratified by smoking status and restricted to individuals of European ancestry. GWAS were stratified by whether people had ever smoked, with never smoked as the comparator. GWAS were conducted using the MRC Integrative Epidemiology Unit UK Biobank GWAS Pipeline (Version 2) [32].

#### Tobacco use

When tobacco use was the exposure, summary-level statistics from the GSCAN GWAS of individuals of European descent were obtained [28]. We have adapted the names of the instruments from the published GWAS (i.e., smoking continuation vs smoking cessation, smoking heaviness vs cigarettes per day) to aid interpretability. We used betas from these summary statistics with UK Biobank excluded to minimise sample overlap and 23&Me removed due to data sharing restrictions.

##### Smoking initiation [SI]

(N = 249,171) refers to a binary phenotype indicating whether an individual has ever smoked regularly, measured by standard deviation change in probability of lifetime regular smoking (∼10-12% increase in the probably of being a regular smoker). The question was assessed in a variety of ways across included cohorts (e.g., “*Have you ever smoked regularly?*”, “*Have you ever smoked >100 cigarettes over the course of your life?*”). The 378 genome-wide significant conditionally independent SNPs associated with smoking initiation explained ∼4% of variance in the trait [28] when tested in an independent sample.

##### Smoking continuation [SC]

(N =143,851) refers to a binary phenotype contrasting current versus former smokers, measured by standard deviation change in probability of continuing to smoke compared with quitting (∼3-5% increase in the probability of being a current smoker compared with former) and was typically assessed through combinations of questions (e.g., “*Do you currently smoke?*” and “*Have you ever smoked regularly?*”). The 24 genome-wide significant conditionally independent SNPs associated with smoking continuation explained ∼1% of variance in the trait [28] when tested in an independent sample.

##### Smoking heaviness [CPD]

(N = 143,210) refers to a quasi-continuous phenotype, representing average number of cigarettes smoked per day (CPD) either as a current or former smoker measured by standard deviation change in CPD categories (∼2-3 additional cigarettes daily) [24]. Self-reported quantities in cohorts with free-text responses were binned (i.e., 1-5 CPD, 6-15 CPD, 16-25 CPD, 26-35 CPD, 36+ CPD) or pre-defined bins were used when cohorts employed these. The 55 genome-wide significant conditionally independent SNPs associated with smoking heaviness explained ∼4% of variance in the trait when tested in an independent sample [28].

#### Cannabis use

Summary-level statistics from the International Cannabis Consortium (ICC) GWAS of individuals of European descent [29] were used to source data on cannabis initiation. We requested 23andMe summary statistics separately and received these summary statistics from the authors meta-analysed with the remaining cohorts, excluding UK Biobank. Cannabis initiation (CI; N = 57,980) refers to a binary phenotype representing whether an individual has ever tried cannabis, asked in a variety of ways across included cohorts (e.g., “*Have you ever in your life used marijuana?*”, “*Have you ever used marijuana (grass, pot) or hashish (hash, hash oil)?*”) but ultimately reflecting ‘lifetime’ use (i.e., ever use). The 8 genome-wide significant independent SNPs associated with cannabis initiation explained 0.15% of variance in the trait [29].

Summary-level statistics from a GWAS by Levey and colleagues [30] of individuals of European descent were used to source data on CUD, which combined data from the Million Veteran Program, the Psychiatric Genetics Consortium, the Lundbeck Foundation Initiative for Integrative Psychiatric Research and deCODE Genetics. CUD (N = 886,025) refers to a binary phenotype representing whether an individual has CUD, measured in a variety of ways across contributing cohorts (e.g., ICD codes, semi-structured interviews). One contributing cohort, iPSYCH2, adjusted analysis for psychiatric diagnoses (i.e., ADHD, autism spectrum disorder, schizophrenia, bipolar disorder and MDD). Adjustment for MDD presents a potential issue for interpretation of the MR analysis examining CUD as an exposure, as this can introduce bias [33]. As such, summary-statistics with iPSYCH2 excluded (N = 785,635) were requested and provided by the lead author. The analysis identified 22 independent genome-wide significant SNPs [30]; an R^2^_XZ_ for the genome-wide significant SNPs was not reported.

#### Major Depressive Disorder

We included two data sources to assess the outcome of MDD. For the main analysis we conducted a GWAS of MDD using data from UK Biobank, a population-based cohort consisting of ∼500,000 people aged between 37 and 73 years recruited between 2006 and 2010 from across the UK [34]. Participants attended a baseline assessment, and subsets of participants completed repeat assessments including an online mental health questionnaire (MHQ) in 2017 [35]. A detailed description of the study design, participants and quality control (QC) methods have been reported previously [34–36]. MDD cases were identified following a validated approach to defining lifetime major depression in UK Biobank which draws on multiple indicators of depression, including help-seeking, hospital admissions, self-reported diagnosis or anti-depressant use and interview-based measures [37]. Full details are provided in Supplementary Notes **S2** – **S5**.

The full data release contains the cohort of successfully genotyped samples (n = 488,377). Analyses were restricted to individuals of ‘European’ ancestry as defined by an in-house k-means cluster analysis [32]. Standard exclusions including withdrawn consent, mismatch between genetic and reported sex, and putative sex chromosome aneuploidy were applied. Additional information about genotyping and imputation can be found in Supplementary Note **S6**. The GWAS were conducted using the linear mixed model (LMM) association method as implemented in BOLT-LMM (v2.3) [32,38] which accounts for relatedness and population stratification. Models were adjusted for age, sex and genotype array. BOLT-LMM association statistics are on the linear scale, so betas and their corresponding standard errors were transformed to log odds ratios and their corresponding 95% confidence intervals by (μ * (1 - μ)), where μ is the prevalence of depression in the subsample of interest (i.e., total sample, ever smokers, never smokers). Smoking status was categorised using self-reported information on smoking status (Variable ID: 20116). Using this variable an ‘ever smokers’ category was derived, defined as currently or previously smoking occasionally, most days or daily (i.e., more than once or twice). Individuals who reported trying smoking once or twice, or reported never smoking, were categorised as never smokers. Flowcharts of samples contributing to each GWAS are provided in Supplementary Figures **S1 – 3**. To explore population stratification, SNP-based heritabilities were calculated using linkage-disequilibrium score regression (LDSC v1.0.1) [39,40] and QQ plots were generated (Supplementary Figure 4). Overall, low LD score intercepts and ratios (Supplementary Table **S1**) suggested that the genomic inflation observed in the QQ plots was not driven by population stratification.

As a supplementary analysis a second set of summary data was drawn from a GWAS meta-analysis reported by Howard and colleagues [41]. Due to data sharing restrictions, 23&Me participants (n = 307,354) were removed. The remaining sample consisted of data from 500,199 participants of European ancestry, comprising 170,756 cases and 329,443 controls. Full information on diagnostic assessment of MDD case status is available in the original report, and includes a broad definition of depression (i.e., “*Have you ever seen a GP/psychiatrist for nerves, anxiety, tension or depression?*”) in UK Biobank [41] and varied assessments of MDD cases in the Psychiatric Genetics Consortium (PGC) meta-analysis [42] which relied on international consensus criteria with cross-checks from expert reviewers [42].

### Statistical analysis

All MR analyses were conducted using R version 4.3.1 and completed using the *TwoSampleMR, MR-PRESSO, MVMR* and *MendelianRandomization* packages [43–46].

#### Selection of genetic instruments

SNPs that were identified as conditionally independent at the genome-wide significant level of significance (*p <* 5 x 10^-8^) in the published GWAS were further clumped using an LD distance threshold of 500kb and r^2^ < 0.001 to ensure complete independence. All identified SNPs were available in the outcome GWAS of MDD performed in UK Biobank. Where SNPs associated with either exposure used in the MVMR were not available in the other exposure dataset, proxy SNPs were identified with a minimum linkage disequilibrium (LD) R^2^ of 0.8. The data sources were harmonized using the *TwoSampleMR* package, in which we excluded palindromic SNPs which could not be aligned based on their minor allele frequency. For the MVMR, combined exposure datasets (i.e., SNPs for SI and CI, SNPs for SI and CUD) were further clumped (LD R^2^ <0.001, >500kb) to remove overlapping loci and ensure overall independence. Supplementary Note **S7** provides additional detail on the harmonisation, clumping and proxy-searching methods.

#### Statistical power calculation

To calculate post-hoc statistical power of our MR analyses we used an online tool (https://shiny.cnsgenomics.com/mRnd/), based on several parameters including the GWAS sample size, ratio of cases to controls and the proportion of variance in the exposure explained by the genetic instruments (R^2^_XZ_). R^2^_XZ_ was estimated as the pseudo-R^2^ value from a regression of each target exposure on its respective PRS, with and without covariates (e.g., ever smoking in UK Biobank regressed on smoking initiation PRS). See Supplementary Note **S8**.

#### Univariable MR

To estimate the total causal effects of tobacco smoking and cannabis use, our primary estimator was the inverse variance weighted (IVW) method. The IVW statistic is a weighted mean of the ratio estimates (i.e., effect of a SNP on the outcome divided by the effect of a SNP on the exposure), where the weights are the inverse-variances of the ratio estimates [47,48]. The IVW approach assumes that all genetic variants are valid instrumental variables (IVs) [48]. The MR approach relies on three core assumptions regarding the exposure-outcome association; (IV1) the genetic instrument is associated with the exposure (i.e., relevance assumption), (IV2) there are no causes of the genetic instrument which also influence the outcome through mechanisms other than the exposure of interest (i.e., independence assumption), and (IV3) the genetic instrument does not affect the outcome other than through the exposure (i.e., exclusion restriction assumption) [16,17]. Horizontal pleiotropy (i.e., a variant associating with multiple traits) can violate IV3 if the variant associates directly with the outcome or via a confounding factor.

To assess the reliability of the IVW estimates under various assumptions, we conducted sensitivity analyses using MR-Egger [49], weighted median [50], weighted mode [51] and MR-PRESSO methods [46]. MR-Egger assumes a non-zero intercept to account for pleiotropy [48,49]. MR-Egger relies on two assumptions: (i) INstrument Strength Independent of Direct Effect (InSIDE; i.e., pleiotropic effects should not be correlated with instrument strength), and (ii) NO measurement error (NOME) [48,49]. The intercept can be used as a test for the presence of directional horizontal pleiotropy and explore whether the InSIDE assumption is violated. Violations of the NOME assumption were assessed using the I^2^_GX_ statistic, in which values between 0.6-0.9 were corrected with simulation extrapolation (SIMEX) and estimates with values less than 0.6 were not reported [52]. Importantly, MR-Egger has substantially lower statistical power than the IVW method. Weighed median- and mode-based methods fit regressions with greater weight assigned to SNPs with more precise ratio estimates [48]. The weighted-median method can provide reliable causal estimates when <50% of the instrument weight does not satisfy MR assumptions [50], and the weighted-mode method can provide reliable causal estimates provided the largest number of genetic variants (i.e., modal ratio estimand) are contributed by valid SNPs also known as the Zero Modal Pleiotropy Assumption (ZEMPA) [48,51]. As these methods use the median and mode of the distribution, they are still influenced by outliers [48]. MR-PRESSO is an extension of the IVW method, in which SNPs that contribute to heterogeneity above simulated expectations (i.e., outliers) are removed following a “leave-one-out” approach, and significant differences in the causal estimate before and after outlier removal are assessed through a distortion test [46,48].

To assess instrument strength (IV1) we computed the F-statistic, whereby F>10 indicates sufficient instrument strength [53]. Cochran’s Q was computed to assess heterogeneity between SNP-estimates in each instrument, whereby the Q-statistic should be less than the number of SNPs [53]. We additionally performed leave-one-out analyses, repeating IVW after removing each SNP, and generated scatterplots of SNP effect sizes to further assess explore heterogeneity and potential pleiotropic effects [54].

#### Sensitivity analysis

For the univariable MR, Steiger filtering was used to explore reverse causality. Steiger filtering computes the amount of variance each SNP explains in the exposure and outcome variables, which can be used to exclude SNPs which explain more variance in the outcome [55]. We also conducted analyses of the smoking continuation and smoking heaviness instruments among never smokers as a negative control analysis to explore potential bias from horizonal pleiotropy (i.e., effects observed among never smokers could indicate SNPs influencing the outcome directly or via another phenotype, but not via the target exposure) [56].

#### Multivariable MR

The independent effects of smoking initiation, cannabis initiation and CUD were explored using two complementary MVMR methods: MVMR-IVW and MVMR-Egger [24,57]. For MVMR-Egger, results are reported for analyses with SNPs oriented with respect to each exposure of interest [58]. We were not able to include the phenotypes of smoking heaviness or smoking continuation as these were analysed in samples of ever smokers only, whereas the cannabis initiation and CUD phenotypes were not analysed stratified by smoking status. For the MVMR we used the Sanderson-Windmeijer conditional F-statistic (F_TS_) to test whether SNPs were strongly associated with each exposure given the other exposures included in the model, or ‘conditional relevance’ [24], where F_TS_>10 is indicative of sufficient instrument strength [44]. An adaptation of the Cochran’s Q statistic was used to detect heterogeneity among the included SNPs for the analysis, where *Q* estimates should be less than the number of SNPs included in the model to indicate no excessive heterogeneity [44].

## RESULTS

### Instrument strength and heterogeneity

The mean F-statistics for the univariable MR were all ≥10 (Supplementary Table **S2**). The conditional F-statistics for the MVMR models were between 2.85 and 9.14 (Supplementary Table **S3**), indicating that the MVMR estimates may be affected by weak instrument bias. The reduction in F-statistic between the univariable and MVMR is likely due to the high level of correlation between the effect of the SNPs on each exposure in the model, which will lower the power and instrument strength. The direction of weak instrument bias in MVMR can be towards or away from the null [29], limiting conclusions about the impact of this bias on observed estimates.

There was evidence of heterogeneity in the SNP effects across all univariable (Supplementary Table **S4**) and multivariable analyses (Supplementary Table **S5**), with *Q* greater than the number of SNPs included in the model and evidence of heterogenous effects present in the scatterplots (Supplementary Figures **S5** – **S9**). However, MR-Egger intercepts were all null (*p* >0.05), suggesting no clear evidence of bias from directional horizontal pleiotropy (Supplementary Tables **S4** - **5**), and leave-one-out analyses did not indicate effects were driven by any one particular SNP (Supplementary Figures **S10** – **S14**).

### Univariable MR

#### Tobacco

Using 302 SNPs (Supplementary Data **S1**) associated with smoking initiation, there was evidence for a total causal effect of smoking initiation on MDD (OR^IVW^ = 1.34, 95% CI = 1.27-1.42; Figure **1**). The direction of effect was consistent across the MR methods tested (Supplementary Table **S2**), with MR-Egger results not presented due to low I^2^_GX_ values (<0.6; Supplementary Table **S4**). MR-PRESSO detected 11 outliers amongst the included SNPs. There was no substantial difference in the estimate before and after outlier correction (*p* of distortion test = 0.977; Supplementary Table **S4**).

**Figure 1.**
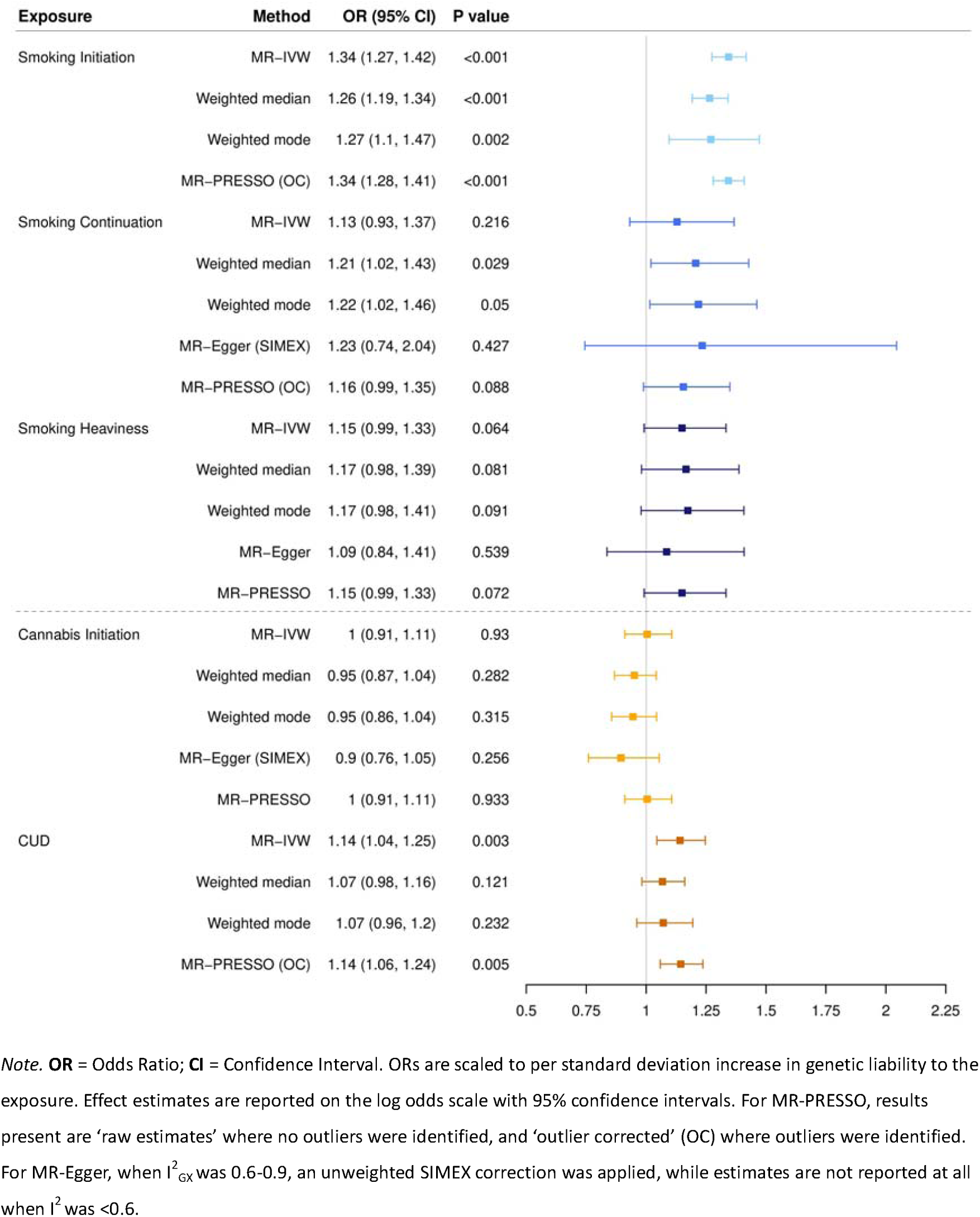
Forest plot depicting univariable MR of the effect of smoking exposures (i.e., initiation, continuation, heaviness), cannabis initiation and CUD on MDD.

Using 17 SNPs (Supplementary Data **S2**) associated with smoking continuation and 40 SNPs (Supplementary Data **S3**) associated with smoking heaviness, univariable MR suggested weaker evidence for total causal effects on MDD (SC: OR^IVW^ = 1.13, 95% CI = 0.93-1.37; CPD: OR^IVW^ = 1.15, 95% CI = 0.99-1.33), with consistent direction of effect across methods (Supplementary Table **S2**; Figure **1**). MR-Egger results for smoking continuation are less reliable due to low I^2^_GX_ (Supplementary Table **S4**). MR-PRESSO detected one outlier (1/17) amongst the included SNPs for smoking continuation. There was no substantial difference in the estimate before and after outlier correction (*p* of distortion test = 0.777; Supplementary Table **S4**).

#### Cannabis

Using 6 SNPs (Supplementary Data **S4**) associated with cannabis initiation, there was no clear evidence for a total causal effect of cannabis initiation on MDD (OR^IVW^ = 1.00, 95% CI = 0.91-1.11; Figure **1**), with consistent direction of effect across methods (Supplementary Table **S2**. Using 16 SNPs (Supplementary Data **S5**) associated with CUD, there was evidence for a causal effect of CUD on MDD (OR^IVW^ = 1.14, 95% CI = 1.04-1.25; Figure **1**) with consistent direction of effect across methods (Supplementary Table **S2**). For cannabis initiation MR Egger results are less reliable due to low I^2^ and not presented for CUD due to I^2^ < 0.6 (Supplementary Table **S4**). MR-PRESSO detected two outliers (2/16) amongst the included SNPs for CUD. There was no substantial difference between the estimate before and after outlier correction (*p* of distortion test = 0.947; Supplementary Table **S4**).

### Multivariable MR

When genetic variants for smoking initiation and cannabis initiation were simultaneously entered in the MVMR model (Model 1; Figure **2**), the independent effects for both exposures (SI: OR^MVMR-IVW^ = 1.36, 95%CI = 1.27 – 1.45; CI: OR^MVMR-IVW^ = 0.98, 95%CI = 0.91 – 1.05, Supplementary Table **S3**), were similar to effect estimates from the univariable MR. When genetic variants for smoking initiation and CUD were simultaneously entered in the MVMR model (Model 2; Figure **2**), the independent effects of smoking initiation (SI: OR^MVMR-IVW^ = 1.29, 95%CI = 1.21 – 1.38) and CUD (CUD: OR^MVMR-IVW^ = 1.06, 95%CI = 1.01 – 1.11; Supplementary Table **S3**) both attenuated when comparing to effect estimates from the univariable MR. Direction of effects were consistent across MVMR-IVW and MVMR-Egger, although with slightly wider confidence intervals for MVMR-Egger.

**Figure 2.**
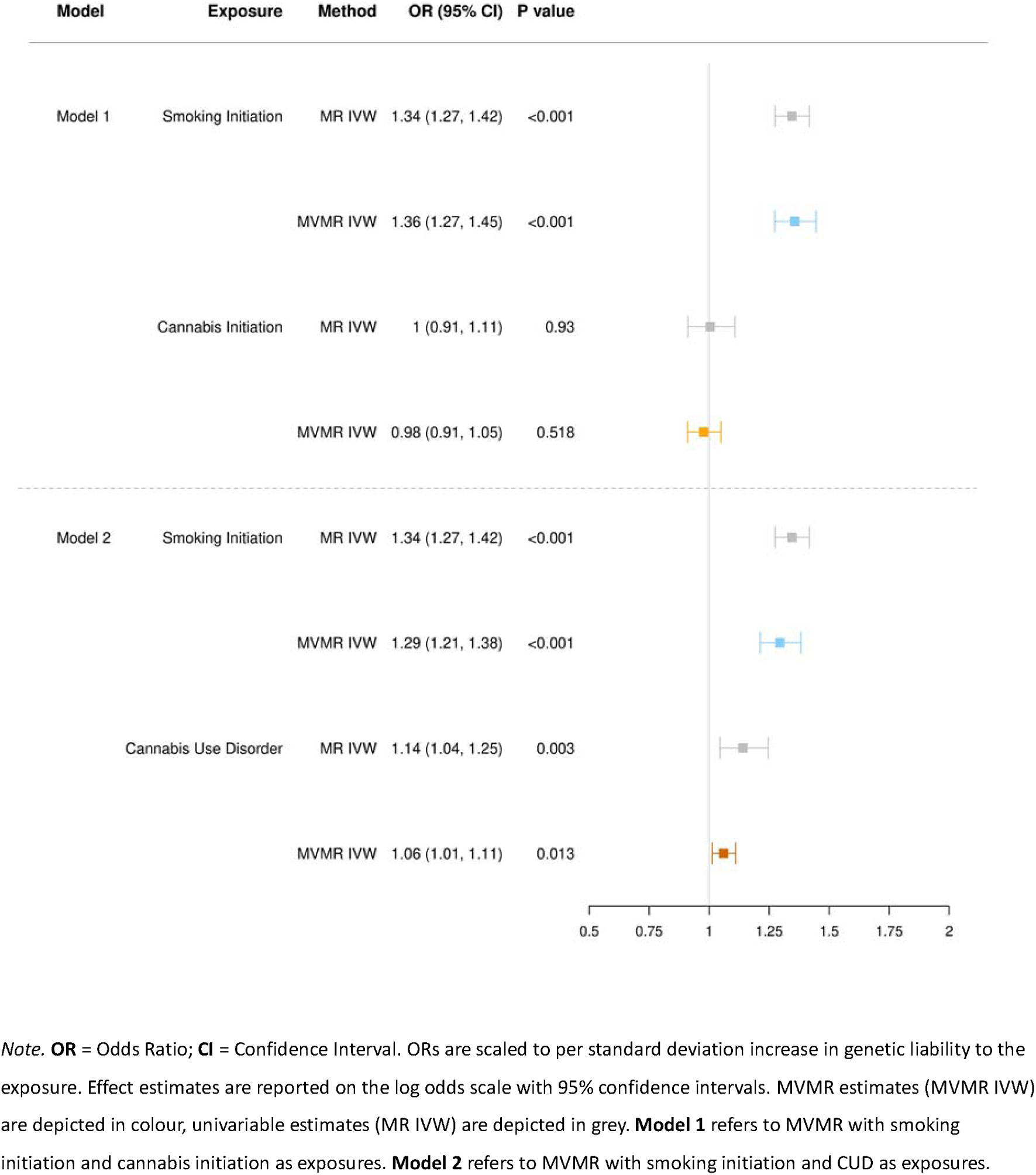
Forest plot comparing univariable and MVMR effects of smoking initiation, cannabis initiation and CUD on MDD.

The conditional F-test demonstrated potential weak instrument bias for all exposures (Model 1: F^TS^ = 9.14 [SI], F^TS^ = 2.85 [CI]; Model 2: F^TS^ = 8.61 [SI], F^TS^ = 4.17 [CUD]), and modified Cochran’s Q statistic indicated evidence of substantial heterogeneity (Model 1: *Q* = 710.88, Model 2: *Q* = 703.62; Supplementary Table **S5**). Results were consistent when heterogeneity tests were repeated with Q-statistic minimisation (Model 1: *Q* = 721.01, Model 2: *Q* = 676.80; Supplementary Table **S5**). Given evidence of weak instruments and heterogeneity, robust estimates were obtained through Q-statistic minimisation using the phenotypic correlation between the exposures (Model 1: *r* = 0.60, Model 2: *r* = 0.61) reported in the published GWAS [24,26]. Results were consistent for the MVMR model of smoking initiation and cannabis initiation on MDD, but the independent effect estimate for CUD on MDD attenuated further towards the null (OR^MVMR-ROB^: OR = 1.03, 95%CI = 0.97 –1.08; Supplementary Table **S3**, Supplementary Figure **S15**).

### Sensitivity analysis

#### Steiger filtering

Steiger filtering identified no SNPs which explained more variance in the outcome for smoking heaviness, cannabis initiation or CUD (Supplementary Table **S6**), indicating limited evidence of reverse causality. Steiger filtering identified 40/302 (13.2%) smoking initiation SNPs which explained more variance in the outcome. Analyses were repeated across methods with these SNPs removed resulting in an attenuated effect (OR^IVW^ = 1.25, 95% CI = 1.20-1.31; Supplementary Table **S6**). Steiger filtering identified 1/17 (5.8%) smoking continuation SNPs for exclusion, and similar effects were observed (OR^IVW^ = 1.16, 95% CI = 0.99-1.35; Supplementary Table **S6**) except for MR-Egger which suggested no clear evidence for a causal effect (OR^EGGER^ = 1.00, 95% CI = 0.64 – 1.57).

#### Negative control analysis

Analyses of smoking continuation and heaviness instruments performed in never smokers indicated no clear evidence of an effect (i.e., supportive of no directional horizontal pleiotropy; Supplementary Table **S7**, Supplementary Figure **S16**).

#### Alternative outcome GWAS

Across the univariable and multivariable MR the effects for smoking initiation, cannabis initiation and CUD were consistent when the summary statistics for MDD were obtained from a GWAS meta-analysis of the Psychiatric Genetics Consortium and UK Biobank (Supplementary Tables **S8**-**9**), with slightly larger effect estimates.

## DISCUSSION

To our knowledge, this is the first study to examine the independent effects of tobacco smoking and cannabis use on MDD using a multivariable MR design. Overall, our results are suggestive of weak evidence that genetic liability to cannabis initiation or CUD has a causal effect on MDD when accounting for the genetic contributions of smoking initiation.

Our findings regarding a causal effect of tobacco smoking for MDD are consistent with previous observational studies demonstrating increased risk of depression amongst individuals who smoke cigarettes [4–6], reduced depressive symptoms following smoking cessation [59] and MR studies suggesting evidence for a causal effect of smoking initiation [18,19,60], lifetime smoking and smoking heaviness [19] on depression. Notably, evidence for a causal effect was weaker when we examined instruments proxying more nuanced phenotypes of smoking behaviour (i.e., smoking continuation, smoking heaviness), where analyses exhibited wider confidence intervals although still consistent evidence regarding direction of effect. It’s possible this relates to the smaller sample sizes contributing to these GWAS, both for the exposure and outcome, which are further restricted to ever smokers where low power will result in imprecise effect estimates [61].

Limited evidence for a causal effect of cannabis initiation on risk of MDD is somewhat inconsistent with previous observational studies, which provide evidence for a slight increase in risk of depression among individuals who have ever used cannabis [9]. However, evidence suggests the frequency and severity of cannabis use is an important moderating factor in the association between cannabis use and depression outcomes with evidence of dose-response effects [62]. Studies employing other genetically informative designs such as twin studies, suggest that robust associations are only observed for higher frequency use rather than dichotomous measures such as ever- or past-year use [63]. Previous research using MR to explore the causal relationship between cannabis initiation and depression has also yielded no clear evidence of causality [20].

When examining the independent causal effects of smoking initiation, cannabis initiation and CUD on MDD using MVMR, our initial results suggested evidence for a causal effect of CUD and smoking initiation independent of one another. However, the conditional F-statistics were all <10, which indicates genetic overlap between these exposures and potential weak instrument bias. This is not unexpected, smoking initiation (i.e., ever regularly smoking) is highly correlated with both cannabis initiation and CUD [28]. Importantly, unlike in univariable MR, weak instrument bias in MVMR can bias the estimated effect of each exposure either towards or away from the null, making it particularly important to test for [44]. Ideally, we would have employed smoking heaviness for this analysis, rather than smoking initiation. However, this would require stratification by smoking status which was not possible using the existing summary statistics for cannabis initiation or CUD.

Due to low instrument strength and high heterogeneity, we re-estimated the MVMR models using weak instrument robust estimators [44]. These results still supported evidence of a causal effect of smoking initiation, but suggested results for CUD were biased away from the null in the primary analysis (OR^MV-ROB^ = 1.03, 95%CI = 0.96 – 1.10; i.e., no clear evidence of a causal effect). These results suggest that tobacco use could, at least in part, underly the associations of CUD with MDD. However, smoking initiation has been found to be horizontally pleiotropic (i.e., associated with numerous other traits) and the attenuation in effect may be capturing other underlying phenotypes (e.g., deprivation, risk-taking) [64], rather than tobacco use specifically.

Furthermore, although simulation analyses have demonstrated that weak instrument robust estimators produce reliable estimates for moderately weak instruments (e.g., F^TS^ =4.23) [44], it is unclear how far this extends and whether this approach generates reliable point estimates for weaker instruments which were employed in our MVMR analysis of cannabis initiation and smoking initiation. As such, our exploration of the independent role of cannabis use, versus tobacco use, on MDD using MVMR should be interpreted with caution.

Another consideration relevant to interpretation of the MVMR is that the lead signal reported in the GWAS of CUD was near *CHRNA2* (rs56372821), which encodes cholinergic receptor nicotinic alpha 2 subunit [30]. This could suggest potential convergence in the cholinergic system and nicotinic receptors in the underlying aetiology of CUD [30]. Results from a multi-trait conditional and joint analysis of CUD and smoking traits (i.e., smoking initiation, smoking heaviness) suggest that 20/22 original lead SNPs remain genome-wide significant after conditioning, and does not substantially alter the magnitude of the lead *CHRNA2* association [30]; a finding replicated in other analyses [65,66]. However, the control for concurrent tobacco use in these analyses is based on estimated rates of tobacco use in which the co-administration of cannabis and tobacco (e.g., in the form of spliffs or blunts) is not captured [67].

Using genetic variants for cannabis initiation is not an ideal measure as it includes those who may have only used cannabis a few times [22]. At the time of conducting the analyses there were not publicly available GWAS of cannabis frequency, although one has recently been published [68]. As such, we employed a genetic instrument for CUD which refers to a pattern of symptoms which cause clinically significant impairment or distress [69] and DSM-5 criteria include experiences of tolerance and unsuccessful reduction or quit attempts [70]. There are no previous MR studies examining the effect of CUD on MDD. Our univariable MR indicated some evidence that CUD has a causal effect on MDD. It is important to highlight that the cannabis instruments employed in the summary-level MR had the lowest power, with genetic instruments explaining only a small proportion of the variance in the target exposure in the UK Biobank sample and with less than 80% power to detect an OR below 1.38 (CI) - 1.55 (CUD). When we replicated analyses using an alternative GWAS, which includes other contributing cohorts besides UK Biobank, effect estimates were slightly larger. However, there is some sample overlap between the exposure and alternative outcome GWAS with a maximum of 0.7% and 9.5% of individuals in cannabis initiation and CUD GWAS, respectively, present in the outcome GWAS; which may bias effect estimates towards the confounded observational estimate (i.e., away from the null) [71]. Notably, results regarding the independent effect of CUD on MDD from the MVMR were consistent when using the alternative outcome GWAS (i.e., no clear evidence of an effect). In summary, due to low power, the presence of small, potentially meaningful, causal effects of CUD on MDD cannot be ruled out and replication in other samples is warranted.

Another consideration relevant to the UK Biobank sample, is the age at which most participants used cannabis. For example, the average age of last cannabis use reported by participants on the UK Biobank data showcase is ∼32 years of age [ID 20455]. Cannabis potency has been reported as an important moderating factor in the association between cannabis use and mental health [72]. A review of studies in this area suggests that the evidence is particularly strong for high potency cannabis and psychosis, whereas evidence for depression is more mixed [72], and highlights that longitudinal observational studies with adjustment for important confounding variables is lacking. Cannabis potency is typically defined by the concentration of Δ^9^-Tetrahydrocannabinol (THC), the primary psychoactive component of cannabis. In Europe, the concentration of THC has more than doubled in the last decade in street cannabis [73,74]. Based on the age of the UK Biobank sample and average age of last use it is likely that, on average, this sample were using a lower potency cannabis than contemporary users. If cannabis potency represents an important moderating factor for the association between cannabis use and depression, replication in studies where participants have been exposed to cannabis that better reflects contemporary cannabis products will be important to understanding health risks.

### Strengths and limitations

This study has numerous strengths including: (i) employing instruments which characterise heaviness of use, (ii) the use of multivariable MR, and (ii) employing a range of sensitivity analyses to support conclusions. However, there are several important limitations to be acknowledged.

MR was employed in this study to minimise the limitations of conventional epidemiological studies. However, MR relies on various assumptions which if violated may generate biased estimates. F-statistics suggested that instruments in the univariable MR were sufficiently associated with the exposure of interest (i.e., IV1), but testing the instruments against the target exposures in UK Biobank suggested weak power to detect the causal effects of cannabis use phenotypes on MDD. Conditional F-statistics suggested the MVMR analyses were limited by weak instruments (F_TS_<10), which we attempted to correct for using robust estimates obtained through Q-minimisation. While the nature of genetic variants being randomly inherited from parents to offspring, means genotypes are typically associated with conventional confounders (e.g., SES) to a much lesser extent (i.e., IV2) [16], population stratification can reintroduce confounding of genotype-outcome associations [75]. All GWAS used to source summary-level data adjusted for population structure in some way (e.g., via adjustment for genetic principal components) and were taken from similar underlying ancestry, However, given that substance use behaviours are socially patterned, residual population structure could reintroduce confounding into the MR analysis [76]. Previous research has reported unmeasured geographical confounding in GWAS of lifestyle-related traits in UK Biobank, even after adjusting for population structure via mixed modelling and principal component analysis [77]. We performed a series of sensitivity analyses to explore potential violation of IV2 (i.e., independence assumption) and IV3 (i.e., exclusion restriction). Several pleiotropy-robust (i.e., MR-Egger, weighted median, weighted mode) methods, outlier-exclusion tests and negative-control analyses, provided limited evidence for violation of these assumptions for the tobacco smoking phenotypes. However, multivariable MR suggested violation due to pleiotropy for the effect of CUD on MDD.

In addition to limitations of the MR approach, there are limitations to the phenotypes and study samples we used in our analyses. First, whilst CUD will reflect heavier frequency of use it is not a direct proxy for cannabis heaviness. CUD affects approximately one in five individuals who use cannabis [69], with higher risk for those who use more frequently (e.g., daily). However, CUD is defined by multiple criteria (e.g., significant impairment in functioning) and is associated with multiple psychological consequences (e.g., financial and social difficulties) [78]. As such, results should be repeated using genetic instruments for cannabis frequency [68], to more accurately estimate the effect of higher frequency cannabis use on depression risk. Second, there is a well-documented selection bias in UK Biobank such that the population are substantially better educated and healthier (e.g., fewer chronic health conditions) which can distort genetic associations and downstream analyses (e.g., MR estimates) especially for socio-behavioural traits [79]. If both cannabis use and MDD influence selection into our study (i.e., less likely to participate) then results may be biased towards the null. Finally, the summary-statistics used to inform the MR analyses were all derived in participants of European ancestry. As such, results may not be generalisable to other populations especially considering the population-specific differences in routes of administration (e.g., in Europe, cannabis is frequently co-administered with tobacco) [12].

## CONCLUSION

The current study provides evidence for a causal effect of tobacco use on risk of MDD. This finding is consistent with evidence from previous observational and MR studies. Our results provide weaker evidence that cannabis initiation or CUD has a causal effect on risk of MDD, independent of liability to smoking initiation. However, we had limited power to detect the causal effect of these phenotypes in UK Biobank. Future studies of the independent effects of these substances should triangulate results with high-quality observational studies which are affected by different underlying sources of bias.

## Acknowledgements

We would like to thank Daniel Levey and his team for their support in provision of CUD summary statistics, and Joëlle Pasman and her team for their support in provision of the lifetime cannabis use summary statistics. Quality Control filtering of the UK Biobank data was conducted by R. Mitchell, G. Hemani, T. Dudding, L. Corbin, S. Harrison, L. Paternoster as described in the published protocol (doi: 10.5523/bris.1ovaau5sxunp2cv8rcy88688v). The MRC IEU UK Biobank GWAS pipeline was developed by B. Elsworth, R. Mitchell, C. Raistrick, L. Paternoster, G. Hemani, T. Gaunt (doi: 10.5523/bris.pnoat8cxo0u52p6ynfaekeigi). This research has been conducted using the UK Biobank Resource under Application Number 9142. We thank all the contributors to the consortia we have used GWAS results from in our analyses. We would also like to thank the research participants and employees of UK Biobank and 23andMe for making this work possible.

## Primary funding

This work is primarily supported by a Society for the Student of Addiction PhD studentship awarded to CB. The funding body had no role the design of the study or collection, analysis, and interpretation of data, or in writing this manuscript.

REW is funded by a postdoctoral fellowship from the South-Eastern Norway Regional Health Authority (2020024). GT is funded by a Cancer Research UK Postdoctoral Fellowship (C56067/A21330). This work was supported by the Medical Research Council (MRC) Integrative Epidemiology Unit at the University of Bristol (MC_UU_00032/07).

## ADDITIONAL INFORMATION

### Contributions

CB, TF, HS, RW and GT conceived the study. CB carried out the data curation and analysis and drafted the initial manuscript. RW and HS provided expert guidance on MR methodology and guided all stages of the analysis. All authors assisted in interpretation of the study results, refining of manuscript drafts and approved the final manuscript.

### Data availability

The data used in this study comes from several sources. Summary statistics for the tobacco use instruments are available at: https://conservancy.umn.edu/items/ca7ed549-636b-41c0-ae79-97c57e266417. The summary statistics for the cannabis initiation instrument, without 23andMe, are available at: https://www.ru.nl/en/bsi. Access to the summary statistics including 23&Me, requires a separate data transfer agreement from 23andMe. Further information about obtaining access to 23andMe are available from: https://research.23andme.com/dataset-access/. The summary statistics for the CUD instrument, including iPSYCH, are available at: https://medicine.yale.edu/lab/gelernter/stats/. Access to the summary statistics excluding iPSYCH was obtained through request to the authors. Primary data from the UK Biobank resource used to derive the summary statistics for the outcome GWAS of MDD are accessible upon application (https://www.ukbiobank.ac.uk/). The PGC MDD summary statistics are available at: https://pgc.unc.edu/for-researchers/download-results/.

### Code availability

A copy of the code used in the MR analyses is available at: https://github.com/chloeeburke/cantob_mvmr.

### Patient and public involvement

Details of public and patient involvement in UK Biobank regarding the MHQ development are available online [35]. For this study there was no specific patient involvement processes in setting the research question, exposures, outcome measures or study methodologies. UK Biobank will disseminate key findings from projects on its website, and we aim to disseminate findings through non-academic platforms including the use of research group partnerships and appropriate social media platforms.

