## Supplementary Methods for "Examining the independent roles of cannabis use and tobacco use in depression risk: a multivariable Mendelian randomisation study"

### SUPPLEMENTARY MATERIALS

### Supplementary Note S1. Deviations from pre-registered analysis plan

An analysis was pre-registered on the Open Science Framework (OSF) on May 3rd 2023 (https://osf.io/bg9vk/). Protocol deviations are reported and discussed below.

**(1) Triangulation with observational analysis and individual-level MR:**

We stated in the pre-registration that we would perform an observational analysis in UK Biobank to examine the association between tobacco use and cannabis use and incident major depressive disorder (MDD), adjusting for a range of confounding variables including polygenic risk scores (PRS). We stated this would be triangulated with findings from analyses using individual-level MR and summary-level MR. Due to expiring data licenses associated with the UK Biobank project (Application ID: 9142), the decision was made to not complete these analyses and instead focus on analysis using the summary-level data generated through performing the GWAS of MDD in UK Biobank.

**(2) Sample for GWAS of MDD in UK Biobank:**

We stated in the pre-registration that we would: (i) restrict to individuals of White British ancestry, and (ii) exclude people from the UK Biobank sample that were related; which we did not do when performing the GWAS of MDD. We pre-specified that our approach to performing the GWAS would be the MRC IEU UK Biobank GWAS pipeline (version 2) (1). This pipeline offers two options to performing GWAS, using either PLINK or BOLT-LMM software, but we did not specify which option we would use. We chose to employ BOLT-LMM, which uses a linear mixed model (LMM) to account for both relatedness and population stratification and allowed for a wider range of individuals to be included in terms of relatedness and ancestry (1,2). Given that we were running GWAS stratified by smoking status (i.e., reduced sample size) it was more advantageous to use a method which preserved a greater sample size. As such, while not excluding related individuals is technically a deviation from our pre-registered protocol, relatedness and population stratification is still accounted for in our GWAS of MDD due to the use of BOLT-LMM.

**(3) Univariable MR methods:**

We stated in the pre-registration that we would use the following MR methods for the univariable MR: inverse-variance weighted (IVW), MR-Egger, weighted median, weighted mode and generalised summary-data based MR (GSMR). We decided to replace GSMR with MR-PRESSO. MR-PRESSO is an extension to the IVW method, which attempts to perform the same type of outlier removal as in the GSMR method (3). By using MR-PRESSO we are still drawing from a range of univariable MR methods which make different underlying assumptions about instrument validity (3).

**(4) Relaxing significance thresholds for SNPs in MVMR:**

We stated in the pre-registration that if there are less than 10 SNPs at genome-wide significance level in the MVMR analysis that we would relax the significance threshold in steps (e.g., $p < 5x10^{-7}$) until 10 SNPs are identified, to a minimum of $p<5x10^{-6}$. Due to the risk of introducing more pleiotropic effects, we instead decided to use a weak-instrument robust version of MVMR (4).

**Supplementary Note S2.** Approach to defining MDD case/control status for GWAS in UK Biobank

To perform the GWAS of MDD in UK Biobank, we followed the approach outlined by Glanville et al., (5) which aims to improve the identification of MDD in UK Biobank using multiple indicators of depression.

We first split the UK Biobank cohort by MHQ participation. For individuals who did not participate in the MHQ, endorsement of five depression phenotypes was calculated ('Help-seeking', 'Self-reported depression', 'Antidepressant usage', 'Depression (Smith)' or 'Hospital (ICD-10) depression'. We then counted the number of measures endorsed by each individual. Detailed information on each phenotype is reported in Supplementary Note **S3**. Cases were defined as individuals endorsing ≥2 depression phenotypes, as the strength of genetic contribution to cases with at least two measures was found to approximate that for CIDI-defined (i.e., gold-standard measure) cases (5).

For individuals who did participate in the MHQ, cases were identified as individuals with a lifetime history of depression from responses to the CIDI depression module. Scoring criteria have been previously defined (6) and are equivalent to the DSM criteria for MDD (6). Detailed information on the items contributing to the CIDI depression module are reported in Supplementary Note **S3**.

Participants were also screened for five psychosis phenotypes: 'Self-reported psychosis', 'Antipsychotic usage', 'Bipolar (Smith)', 'Hospital (ICD-10) psychosis' and 'Psychosis (MHQ screen)'. Detailed information on these phenotypes are reported in Supplementary Note **S4**. Individuals meeting criteria for any of the psychosis phenotypes were excluded from the analysis (i.e., did not meet case or control criteria). Controls comprised all UK Biobank participants who did not meet the criteria for depression or psychosis phenotypes, as follows:

**(1)** Did not meet the criteria for any indication of depression i.e., 'Help-seeking', 'Self-reported depression', 'Antidepressant usage', 'Depression (Smith), 'Hospital (ICD-10) depression' or 'Lifetime depression (MHQ)'; **AND**

**(2)** Did not meet the criteria for any indication of psychosis i.e., 'Self-reported psychosis', 'Antipsychotic usage', 'Bipolar (Smith), 'Hospital (ICD-10) psychosis' or 'Psychosis (MHQ)'; **AND**

**(3)** Did not endorse the question: "Have you been diagnosed with one or more of the following mental health problems by a professional, even if you do not have it currently [UK Biobank Field ID 20544] for depression (11 = "Depression") in the MHQ Section A screening questions.

Supplementary Note S3. Derivation of depression phenotypes in UK Biobank

This describes the criteria for defining cases for each of the depression phenotypes derived from different sources of phenotypic information in UK Biobank. [ID] refers to the corresponding UK Biobank Field ID, which are browsable via the data showcase platform: https://biobank.ndph.ox.ac.uk/showcase/.

**Help-seeking**

Criteria for defining 'help-seeking' cases was endorsing either of the following questions at baseline, or the subsequent repeat assessments:

| Measure | Coding | ID |
|---|---|---|
| "Have you ever seen a general practitioner (GP) for nerves, anxiety, tension or depression?" | 1 = "Yes" | 2090 |
| "Have you ever seen a psychiatrist for nerves, anxiety, tension or depression?" | 1 = "Yes" | 2100 |

**Self-reported depression**

Criteria for defining 'self-reported depression' was endorsing "depression" at baseline or the subsequent repeat assessments:

| Measure | Coding | ID |
|---|---|---|
| Code for non-cancer illness. If the participant was uncertain of the type of illness they had had, then they described it to the interviewer (a trained nurse) who attempted to place it within the coding tree. If the illness could not be located in the coding tree then the interviewer entered a free-text description of it. These free-text descriptions were subsequently examined by a doctor and, where possible, matched to entries in the coding tree. Free-text descriptions which could not be matched with very high probability have been marked as "unclassifiable". | 1286 = "depression" | 20002 |

**Antidepressant usage**

Criteria for defining 'antidepressant usage' cases was self-reported antidepressant medication at the baseline or the subsequent repeat assessments:

| Measure | Codes | ID |
|---|---|---|
| Medication Status was obtained via a verbal interview item requesting the names of regular prescription medications that the participants were currently taking. The nurses conducting the interviews did not record medications that were short-term (e.g., 1-week course of antibiotics), historical, or prescribed but not being taken. | 1140879616, 1140921600, 1140879540, 1140867878, 1140916282, 1140909806, 1140867888, 1141152732, 1141180212, 1140879634, 1140867876, 1140882236, 1141190158, 1141200564, 1140867726, 1140879620, 1140867818, 1140879630, 1140879628, 1141151946, 1140867948, 1140867624, 1140867756, 1140867884, 1141151978, 1141152736, 1141201834, 1140867690, 1140867640, 1140867920, 1140867850, 1140879544, 1141200570, 1140867934, 1140867758, 1140867914, 1140867820, 1141151982, 1140882244, 1140879556, 1140867852, 1140867860, 1140917460, 1140867938, 1140867856, 1140867922, 1140910820, 1140882312, 1140867944, 1140867784, 1140867812, 1140867668 | 20003 |

**Depression (Smith)**

Criteria for defining ‘Depression (Smith)’ cases was meeting the criteria for one of three depression phenotypes at baseline, defined previously by Smith et al., (7) and described in detail under resource 158722:

| **Measure** | **Codes** | **ID** |
| --- | --- | --- |
| Single Probable major depression episode (7) | 5 = Single Probable major depression episode | 20126 |
| Probable recurrent major depression (moderate) (7) | 4 = Probable recurrent major depression (moderate) | 20126 |
| Probable Recurrent major depression (severe) (7) | 3 = Probable Recurrent major depression (severe) | 20126 |

**Hospital (ICD-10) depression**

Criteria for ‘Hospital (ICD-10)’ cases was being admitted for hospital inpatient care with a diagnosis (either primary or secondary) for major depression ([ICD-10] = F32.X and F33.X) between April 1997 and November 2023:

| **Measure** | **Codes** | **ID** |
| --- | --- | --- |
| Depressive episode | F32, F320, F321, F322, F323, F328, F329 | 41202<br>41204 |
| Recurrent depressive disorder | F33, F330, F331, F332, F334, F338, F339 | 41202<br>41204 |

**Lifetime depression (MHQ)**

Criteria for defining ‘Lifetime Depression (MHQ) cases is detailed below. These mirror criteria defined by Davis et al., (6) for identifying individuals with a lifetime history of depression, and mirrors CIDI-SF criteria for lifetime depression. Participants must have endorsed: (i) at least one of the two ‘core symptoms’; **AND** (ii) a score above threshold on the ‘threshold items’; **AND** (iii) experiencing ≥5 symptoms (including core) during ‘worst episode of depression’:

| **Measure** | **Codes** | **ID** |
| --- | --- | --- |
| Core symptoms: | 1 = “Yes” | 20446<br>20441 |

| | | |
|---|---|---|
| • "Have you ever had a time in your life when you felt sad, blue, or depressed for two weeks or more in a row?";<br>• "Have you ever had a time in your life lasting two weeks or more when you lost interest in most things like hobbies, work, or activities that usually give you pleasure?" | | |
| Threshold items:<br>"Please think of the two-week period in your life when your feelings of depression or loss of interest were the worst."<br>• "How much of the day did these feelings usually last?" (>2);<br>• "(How often) did you feel this way?" (>1)<br>• "Think about your roles at the time of this episode, including study/employment, childcare and housework, leisure pursuits. How much did these problems interfere with your life or activites?" (>1) | 2 = "About half of the day"<br>1 = "Less often"<br>1 = "A little" | 20436<br>20439<br>20440 |
| Symptoms during worst episode of depression:<br>"Please think of the two-week period in your life when your feelings of depression or loss of interest were the worst"<br>• "Did you feel more tired out or low on energy than is usual for you?"<br>• "Did you gain or lose weight without trying or did you stay about the same weight?"<br>• "Did your sleep change?"<br>• "Did you have a lot more trouble concentrating than usual?"<br>• "People sometimes feel down on themselves, no good, worthless. Did you feel this way?" | 1 = "Yes"<br>1 = "Gained weight"; 2 = "Lost weight"; 3 = "Both gained and lost some weight"<br>1 = "Yes"<br>1 = "Yes"<br>1 = "Yes" | 20449<br>20536<br>20532<br>20435<br>20450<br>20437 |

| | |
|---|---|
| • “Did you think a lot about death – either your own, someone else’s or death in general?” | 1 = “Yes” |

Supplementary Note S4. Derivation of psychosis phenotypes in UK Biobank

This describes the criteria for defining cases for each of the psychosis phenotypes derived from different sources of phenotypic information in UK Biobank. [ID] refers to the corresponding UK Biobank Field ID, which are browsable via the data showcase platform: https://biobank.ndph.ox.ac.uk/showcase/.

**Self-reported psychosis**

Criteria for defining 'self-reported psychosis was endorsing "schizophrenia" or "mania/bipolar disorder/manic depression" at baseline or the subsequent repeat assessments:

| Measure | Coding | ID |
|---|---|---|
| Code for non-cancer illness. If the participant was uncertain of the type of illness they had had, then they described it to the interviewer (a trained nurse) who attempted to place it within the coding tree. If the illness could not be located in the coding tree then the interviewer entered a free-text description of it. These free-text descriptions were subsequently examined by a doctor and, where possible, matched to entries in the coding tree. Free-text descriptions which could not be matched with very high probability have been marked as "unclassifiable". | 1289 = "schizophrenia"<br>1291 = "mania/bipolar disorder/manic depression" | 20002 |

**Antipsychotic usage**

Criteria for defining 'antidepressant usage' cases was self-reported antipsychotic medication at the baseline or the subsequent repeat assessments:

| Measure | Codes | ID |
|---|---|---|
| Medication Status was obtained via a verbal interview item requesting the names of regular prescription medications that the participants were currently taking. The nurses conducting the interviews did not record medications that were | 1140868170, 1140928916, 1141152848, 1140867444, 1140879658, 1140868120, 1141153490, 1140867304, 1141152860, 1140867168, | 20003 |

| | |
|---|---|
| short-term (e.g., 1-week course of antibiotics), historical, or prescribed but not being taken. | 1141195974, 1140867244, 1140867152, 1140909800, 1140867420, 1140879746, 1141177762, 1140867456, 1140867952, 1140867150, 1141167976, 1140882100, 1140867342, 1140863416, 1141202024, 1140882098, 1140867184, 1140867092, 1140882320, 1140910358, 1140867208, 1140909802, 1140867134, 1140867306, 1140867210, 1140867398, 1140867078, 1140867218, 1141201792, 1141200458, 1140867136, 1140879750, 1140867180, 1140867546, 1140928260, 1140927956 |

**Bipolar (Smith)**

Criteria for defining 'Bipolar (Smith)' cases was meeting the criteria for one of two bipolar phenotypes at baseline, defined previously by Smith et al., (7) and described in detail under resource 158722:

| Measure | Codes | ID |
|---|---|---|
| Bipolar Type I (Mania) (7) | 1 = Bipolar Type I (Mania) | 20126 |
| Bipolar Type II (Hypomania) (7) | 2= Bipolar Type II (Hypomania) | 20126 |

**Hospital (ICD-10) psychosis**

Criteria for 'Hospital (ICD-10)' cases was being admitted for hospital inpatient care with a diagnosis (either primary or secondary) for a psychotic disorder between April 1997 and November 2023:

| Measure | Codes | ID |
|---|---|---|

| | | |
|---|---|---|
| Schizophrenia, schizotypal and delusional disorders | F20, F200, F201, F202, F203, F204, F205, F206, F208, F209, F21, F22, F220, F228, F229, F23, F230, F231, F232, F233, F238, F239, F24, F25, F250, F251, F252, F258, F259, F28, F29 | 41202<br>41204 |
| Mood [affective] disorders (excluding Depression codes F32-F33) | F30, F300, F301, F302, F308, F309, F31, F310, F311, F312, F313, F314, F315, F316, F317, F318, F319, F34, F340, F341, F348, F349, F38, F380, F381, F388, F39 | 41202<br>41204 |

**Psychosis (MHQ)**

Criteria for 'Psychosis (MHQ)' cases was endorsing "schizophrenia", "any other type of psychosis or psychotic illness" or "mania/hypomania/bipolar/manic-depression" in response to a screening question in MHQ Section A:

| **Measure** | **Codes** | **ID** |
|---|---|---|
| "Have you been diagnosed with one or more of the following mental health problems by a professional, even if you don't have it currently?" | 2 = "Schizophrenia"<br>3 = "Any other type of psychosis or psychotic illness"<br>10 = "Mania, hypomania, bipolar or manic-depression" | 20544 |

### Supplementary Note S6. Information about the genotyping, quality control and imputation methods for UK Biobank data

This information is taken from the MRC IEU UK Biobank GWAS pipeline (Version 2) recommended paragraphs for publication (1).

The full data release contains the cohort of successfully genotyped samples (n=488,377). 49,979 individuals were genotyped using the UK BiLEVE array and 438,398 using the UK Biobank axiom array. Pre-imputation QC, phasing and imputation are described elsewhere (8). In brief, prior to phasing, multiallelic SNPs or those with MAF ≤1% were removed. Phasing of genotype data was performed using a modified version of the SHAPEIT2 algorithm (9). Genotype imputation to a reference set combining the UK10K haplotype and HRC reference panels (10) was performed using IMPUTE2 algorithms (11). The analyses presented here were restricted to autosomal variants within the HRC site list using a graded filtering with varying imputation quality for different allele frequency ranges. Therefore, rarer genetic variants are required to have a higher imputation INFO score (Info>0.3 for MAF >3%; Info>0.6 for MAF 1-3%; Info>0.8 for MAF 0.5-1%; Info>0.9 for MAF 0.1-0.5%) with MAF and Info scores having been recalculated on an in-house derived 'European' subset (8).

Individuals with sex-mismatch (derived by comparing genetic sex and reported sex) or individuals with sex-chromosome aneuploidy were excluded from the analysis (n=814). We restricted the sample to individuals of 'European' ancestry as defined by an in-house k-means cluster analysis performed using the first 4 principal components provided by UK Biobank in the statistical software environment R. The current analysis includes the largest cluster from this analysis (n=464,708) (8). To model population structure in the sample we used 143,006 directly genotyped SNPs, obtained after filtering on MAF > 0.01; genotyping rate > 0.015; Hardy-Weinberg equilibrium p-value < 0.0001 and LD pruning to an $r^2$ threshold of 0.1 using PLINKv2.00.

### Supplementary Note S7. Additional details regarding harmonisation and clumping

We used the *TwoSampleMR* R package (12) *harmonise_data* function to harmonise the exposure and outcome datasets. Palindromic SNPs were only excluded if their allele frequency could not be used to infer which strand was positive (i.e., action =2). The effect allele frequency (EAF) and minor allele frequency (MAF) were not available in the summary-level statistics provided for the CUD GWAS, due to data sharing restrictions. The lead author informed us that the MAFs were highly similar to those obtainable from the 1000 Genomes Project, and we therefore imputed EAF based on this data as is applied in other GWAS (e.g., GSCAN) (13).

All SNPs available in the exposure GWAS datasets were available in the MDD (UK Biobank) GWAS dataset. There were some SNPs (SI nSNPs = 2, CI nSNPS = 1) available in the exposure GWAS datasets that were not available in the MDD (Howard) GWAS dataset. Due to the low number of SNPs, and as this was a supplementary analysis, we did not search for missing SNPs. For the MVMR, there was one SNP in the cannabis initiation instrument (rs9773390) that was missing from the smoking initiation summary statistics, and one SNP in the smoking initiation instrument (rs2359180) that was missing from the cannabis initiation summary statistics. We used the *LDproxy* function from the LDlinkR package (14) to identify proxy SNPs with a minimum linkage disequilibrium ($R^2$) of 0.8.

In MVMR analyses, all SNPs included in the model should be independent of each other (i.e., SNPs associated with NMR must also be independent of the SNPs associated with CPD, and vice versa) (15). To ensure overall independence, the full list of SNPs were clumped (LD $R^2 < 0.001$, >500kb). Considering the limited number of SNPs associated with both cannabis phenotypes, compare to the smoking initiation instrument, SNPs associated with smoking initiation were dropped from the analysis to preserve instrument strength (i.e., no cannabis initiation or CUD SNPs were dropped during the clumping stage).

Supplementary Note S8. Power analysis for summary-level univariable MR

Power calculations were conducted using the online power calculator for Mendelian randomisation (https://shiny.cnsgenomics.com). We input the sample size contributing to each outcome GWAS in UK used in the primary analyses (i.e., full sample, ever smokers). For the variance explained by the instrument, we used pseudo-$R^2$ estimated within the UK Biobank cohort. In the table below, we present the smallest OR per standard deviation of the exposure variable that we have 80% power to detect.

| **Exposure** | **MDD sample** | **N** | **K** | **$R^2_{xz}$** | **OR (80% power)** |
|---|---|---|---|---|---|
| Smoking Initiation | Full sample | 356641 | 0.22652 | 0.0179 | 1.09 |
| Smoking Continuation | Ever smokers | 160248 | 0.25484 | 0.00019 | 1.39 |
| Smoking Heaviness | Ever smokers | 160248 | 0.25484 | 0.0112 | 1.16 |
| Cannabis Initiation | Full sample | 356641 | 0.22652 | 0.0010 | 1.38 |
| Cannabis Use Disorder | Full sample | 356641 | 0.22652 | 0.0005 | 1.55 |

There is no universal rule for interpreting the size of an OR in terms of "small", "medium" or "large". As general rule of thumb (16), assuming a disease rate of 10%, thresholds for small (OR = 1.46), medium (OR = 2.50) and large (OR = 4.14) effect sizes equivalent to Cohen's d (0.2 = small, 0.5 = medium and 0.8 = large) can be approximated. However, the disease rate in the UK Biobank sample was >10% meaning that these approximations may not be valid as they rely on the assumption that the OR provides a reasonable estimate of the RR (i.e., low population rate of cases).

$R^2_{XZ}$ was estimated as the pseudo-$R^2$ value from a regression of each target exposure on its respective PRS (e.g., ever smoking in UK Biobank regressed on smoking initiation PRS). The proportion of variance attributable to the PRS was estimated as the difference between the Nagelkerke's pseudo-$R^2$ for a model with covariates alone (i.e., age, sex, genotype array, first 10 principal components of ancestry) and a model which included covariates and the PRS [i.e., pseudo-$R^2$ (full model) – pseudo-$R^2$ (model without PRS)].

**Supplementary Figure S1.** Flowchart of GWAS of MDD in UK Biobank in unstratified sample

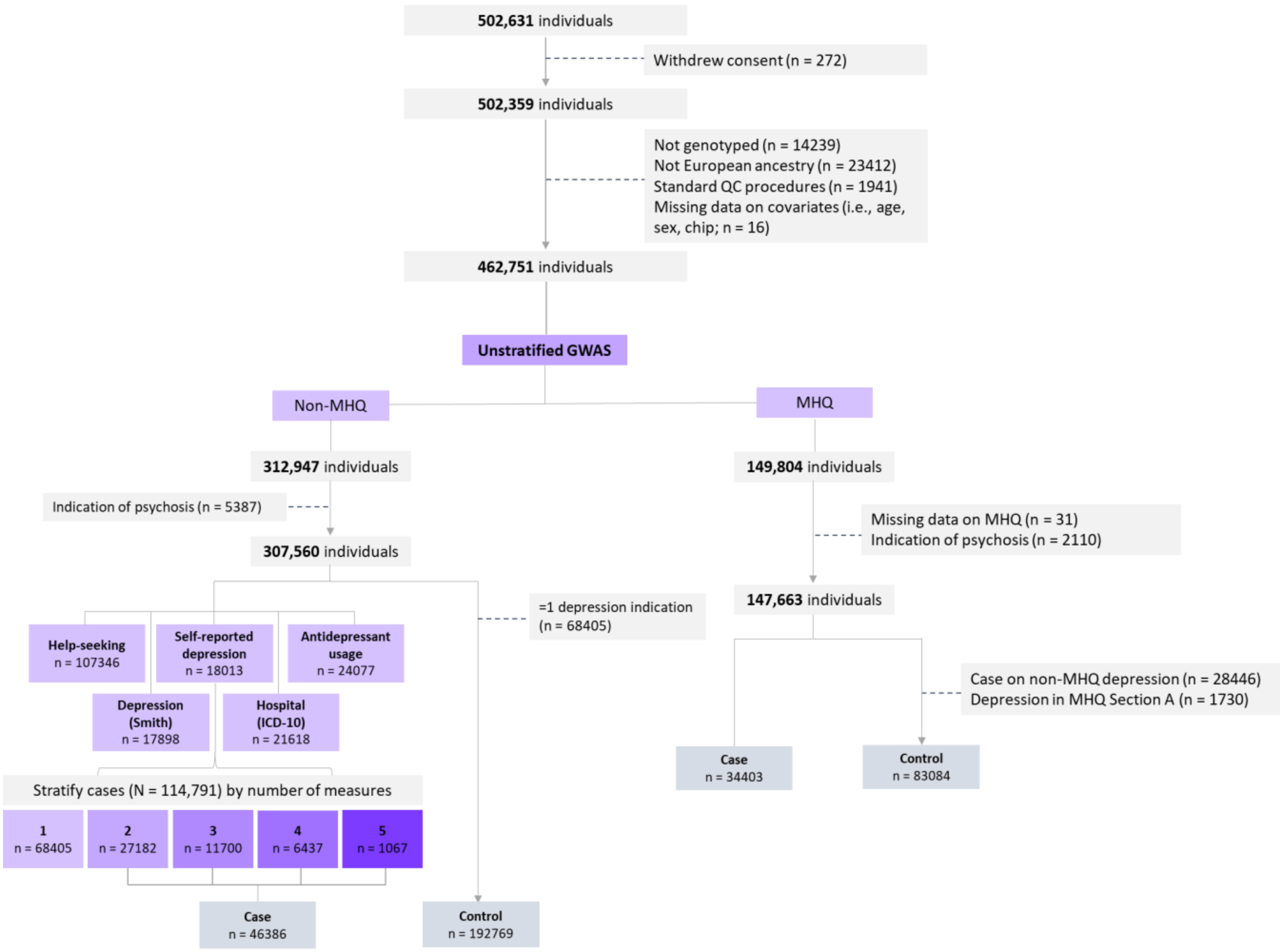

**Supplementary Figure S2.** Flowchart of GWAS of MDD in UK Biobank in ever smokers

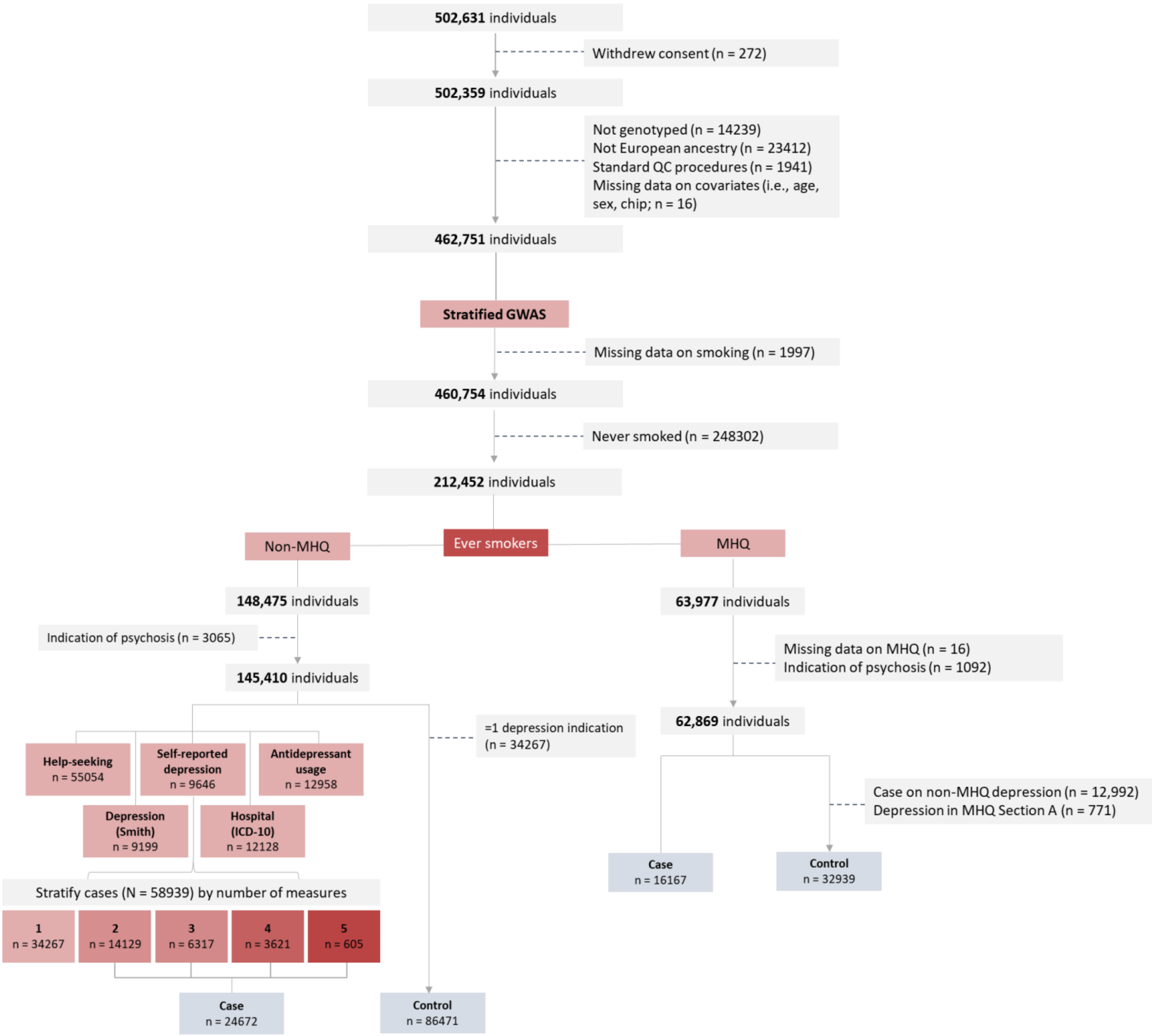

Supplementary Figure S3. Flowchart of GWAS of MDD in UK Biobank in never smokers

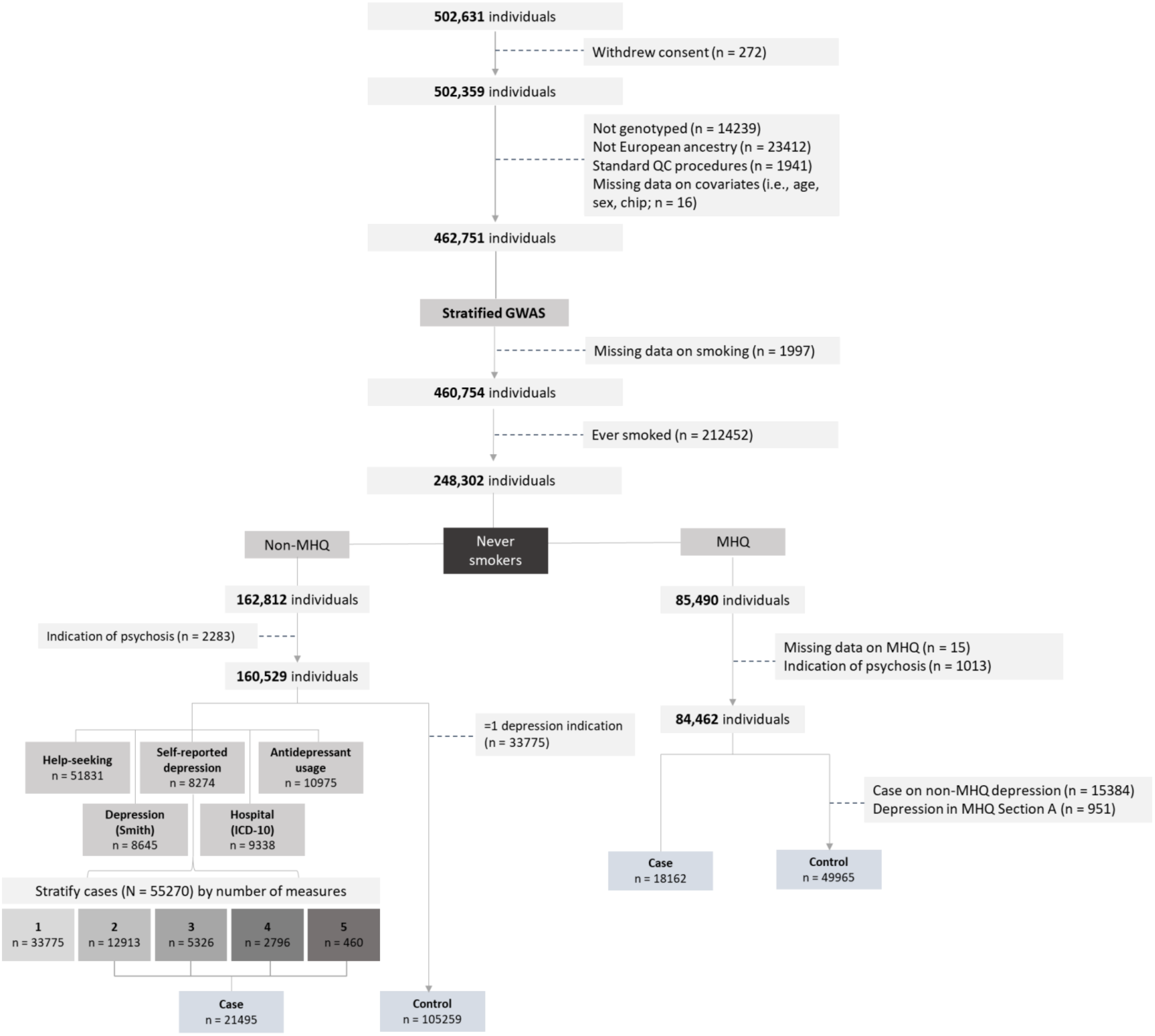

**Supplementary Figure S4.** QQ plots for GWAS of MDD in UK Biobank among (A) full sample, (B) ever smokers, and (C) never smokers

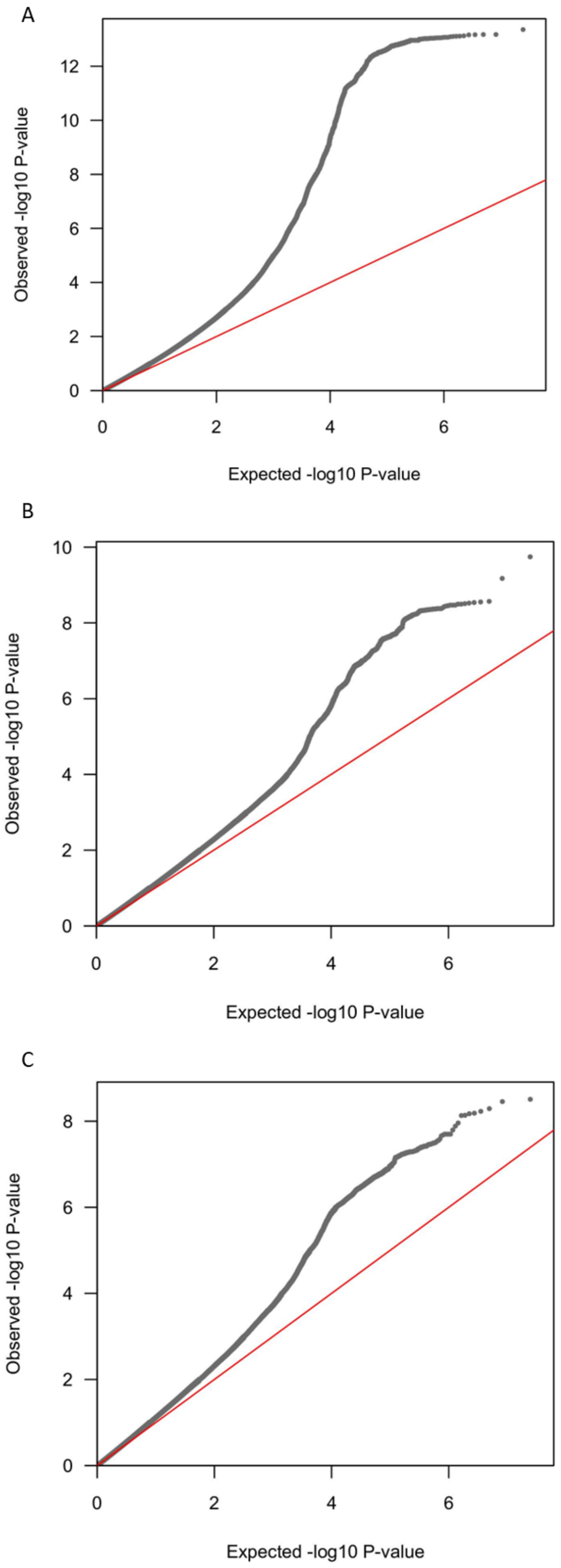

**Supplementary Figure S5.** Scatterplot of the univariable MR analysis of liability to smoking initiation on MDD risk.

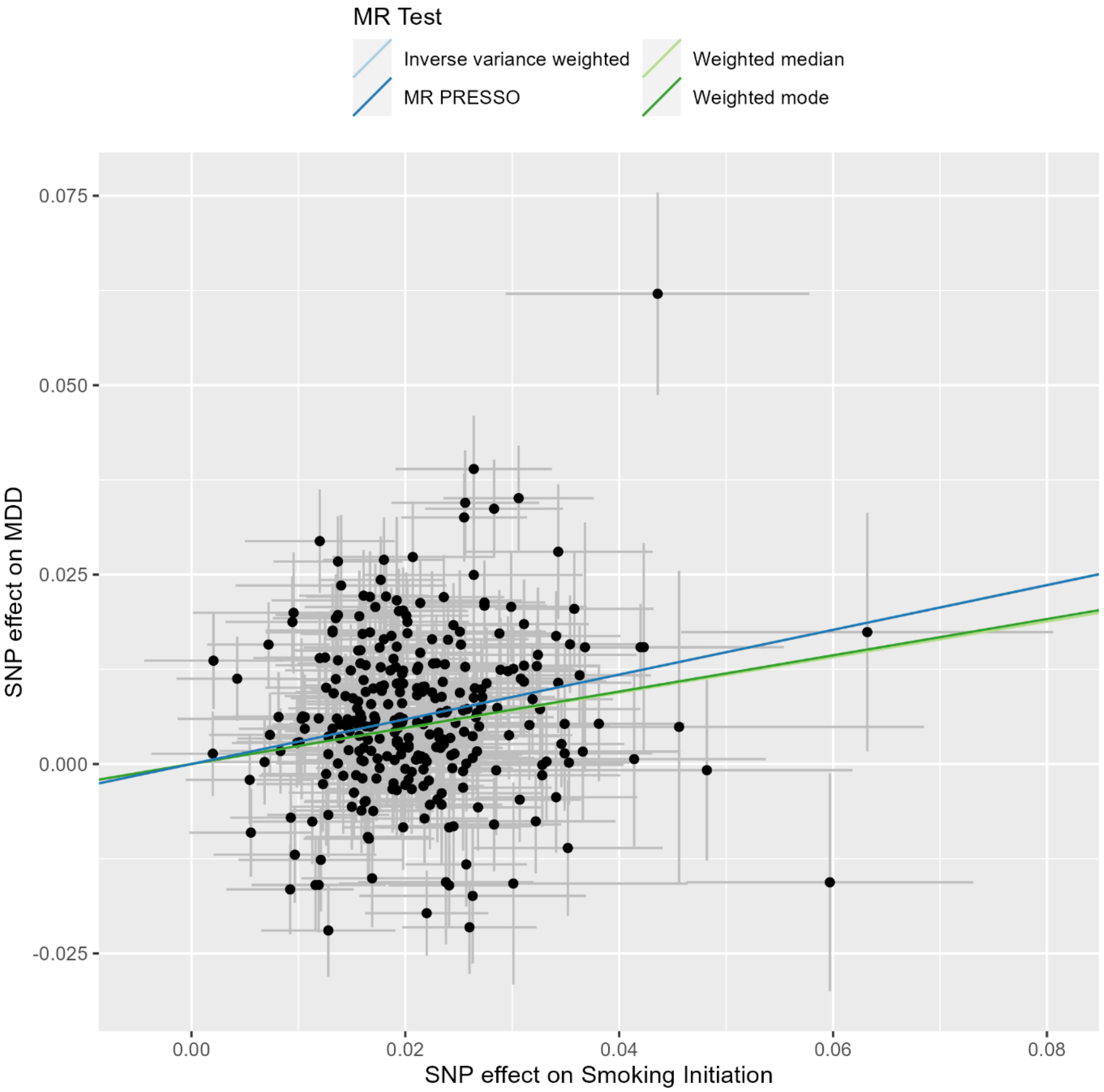

**Supplementary Figure S6.** Scatterplot of the univariable MR analysis of liability to smoking continuation on MDD risk.

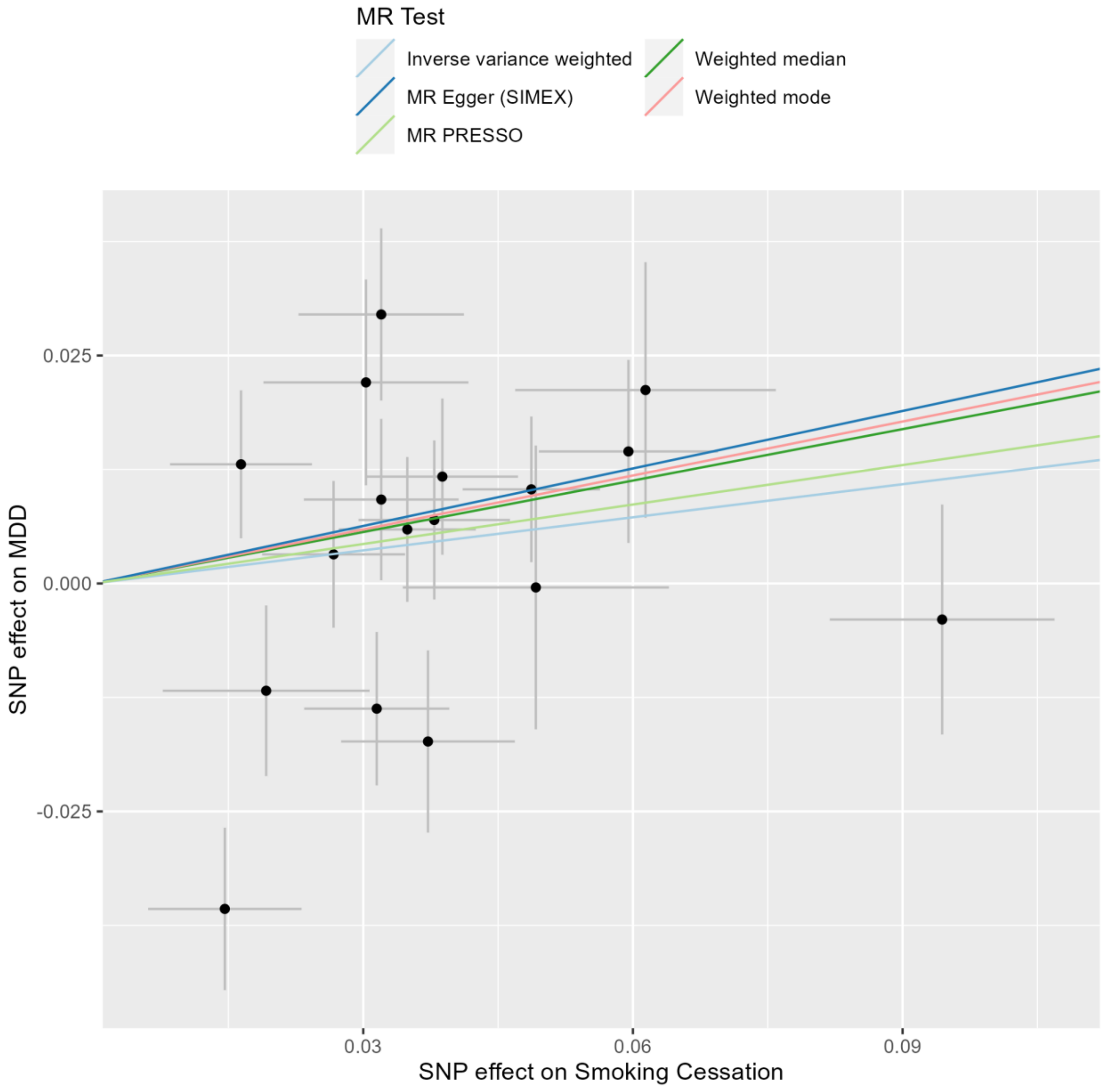

**Supplementary Figure S7.** Scatterplot of the univariable MR analysis of liability to smoking heaviness on MDD risk.

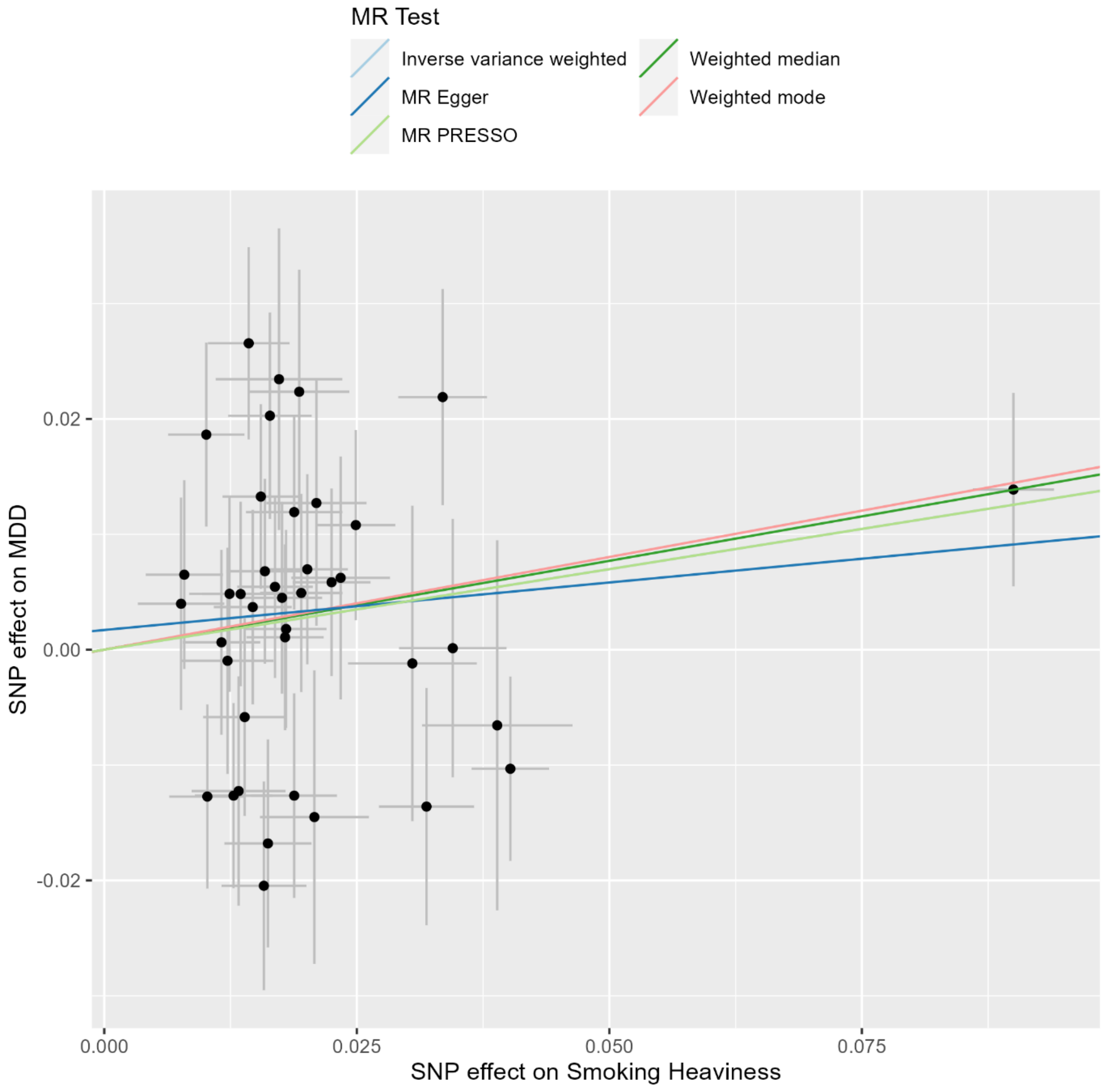

**Supplementary Figure S8.** Scatterplot of the univariable MR analysis of liability to cannabis initiation on MDD risk.

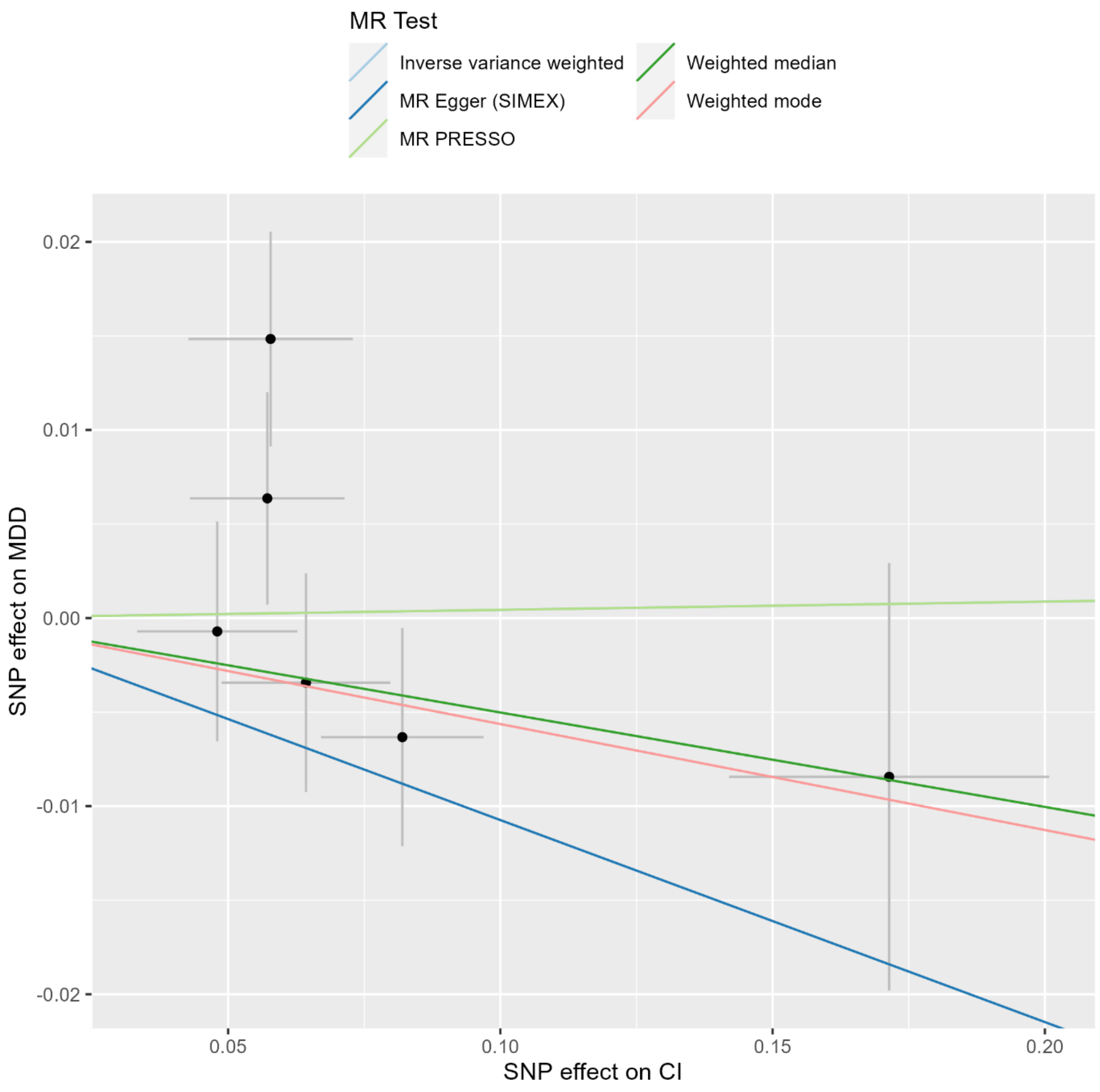

**Supplementary Figure S9.** Scatterplot of the univariable MR analysis of liability to CUD on MDD risk.

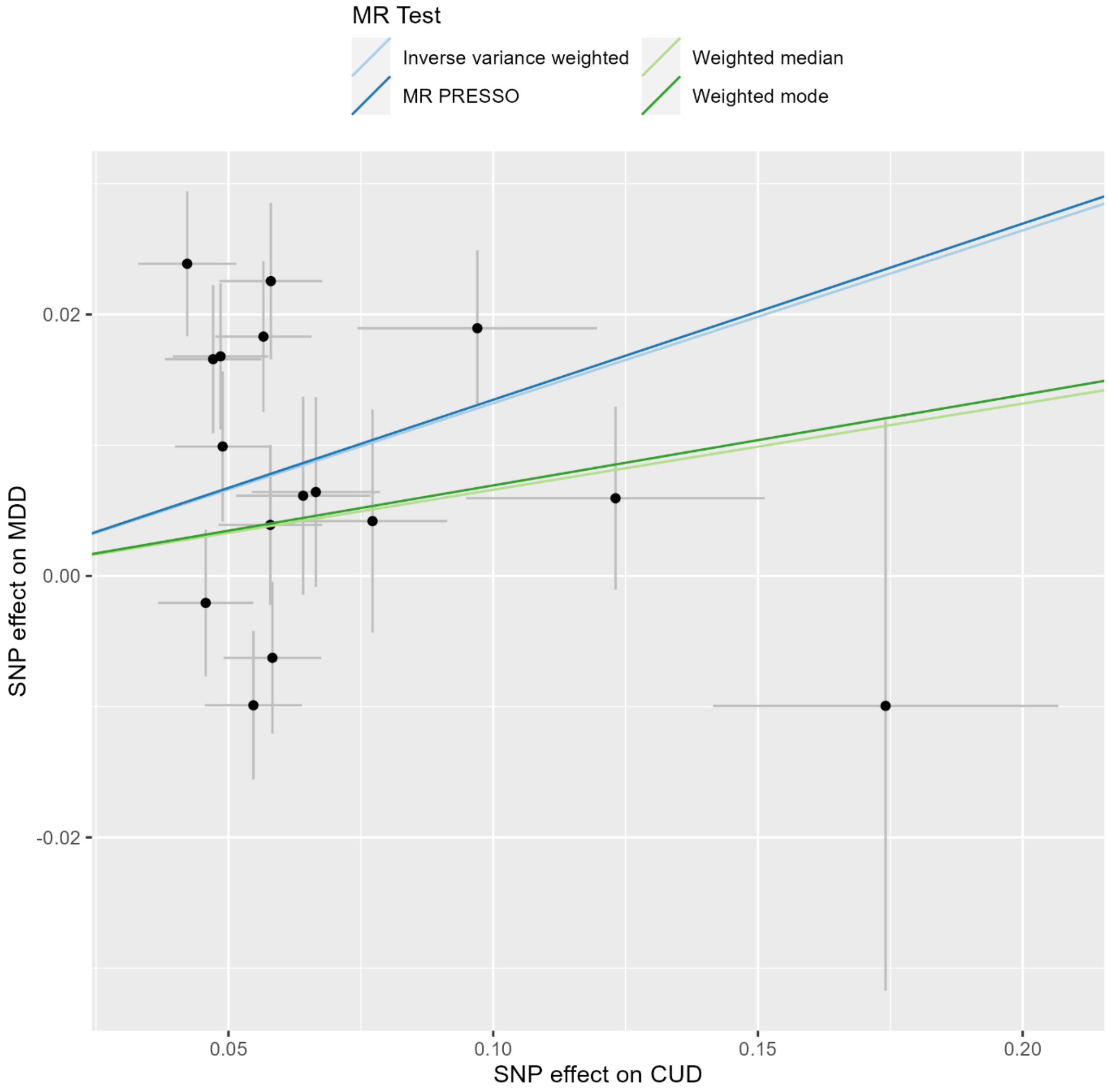

Supplementary Figure 10. Leave-one-out IVW regression analyses of liability to smoking initiation on MDD risk.

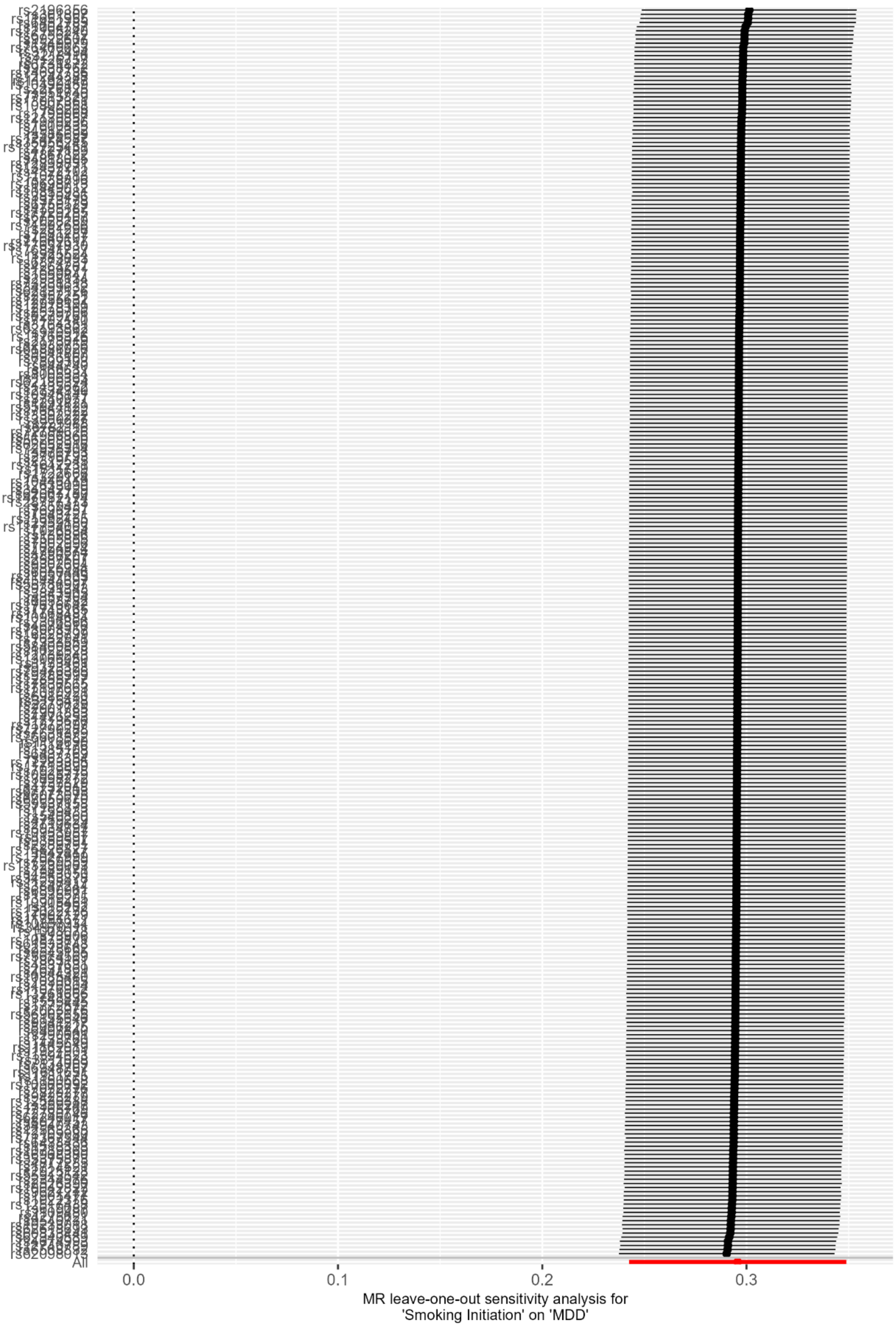

**Supplementary Figure S11.** Leave-one-out IVW regression analyses of liability to smoking continuation on MDD risk.

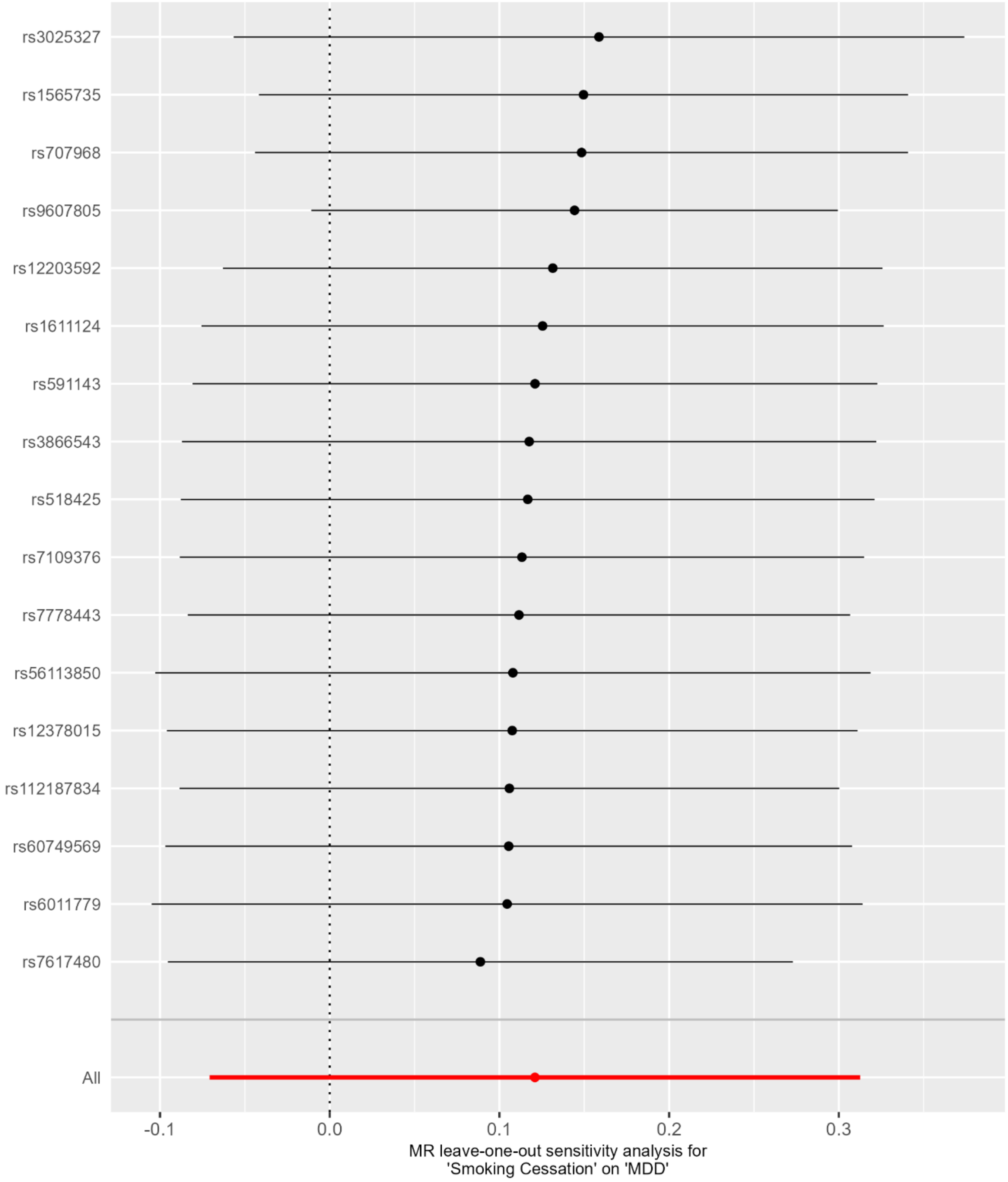

**Supplementary Figure S12.** Leave-one-out IVW regression analyses of liability to smoking heaviness on MDD risk.

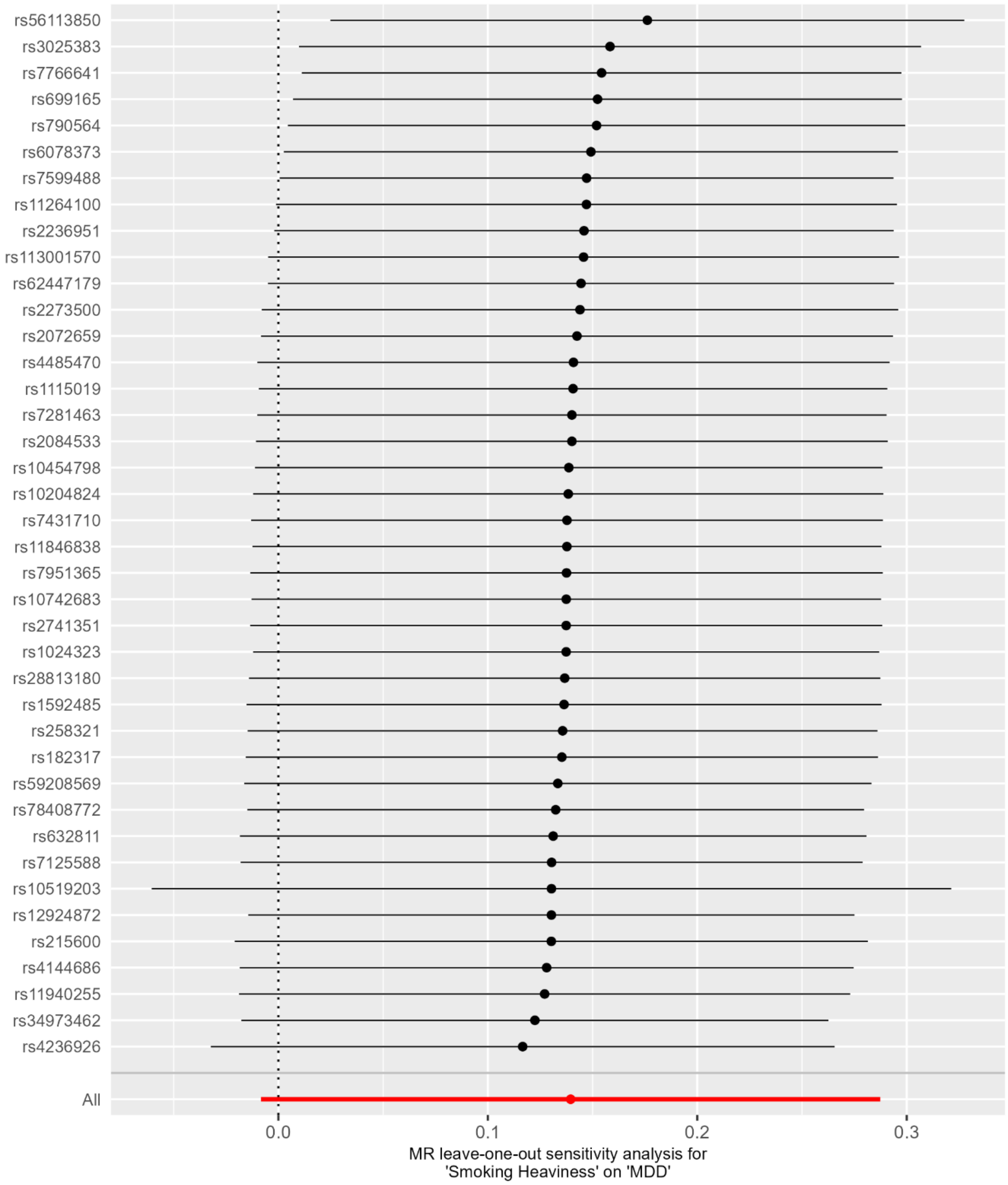

**Supplementary Figure S13.** Leave-one-out IVW regression analyses of liability to cannabis initiation on MDD risk.

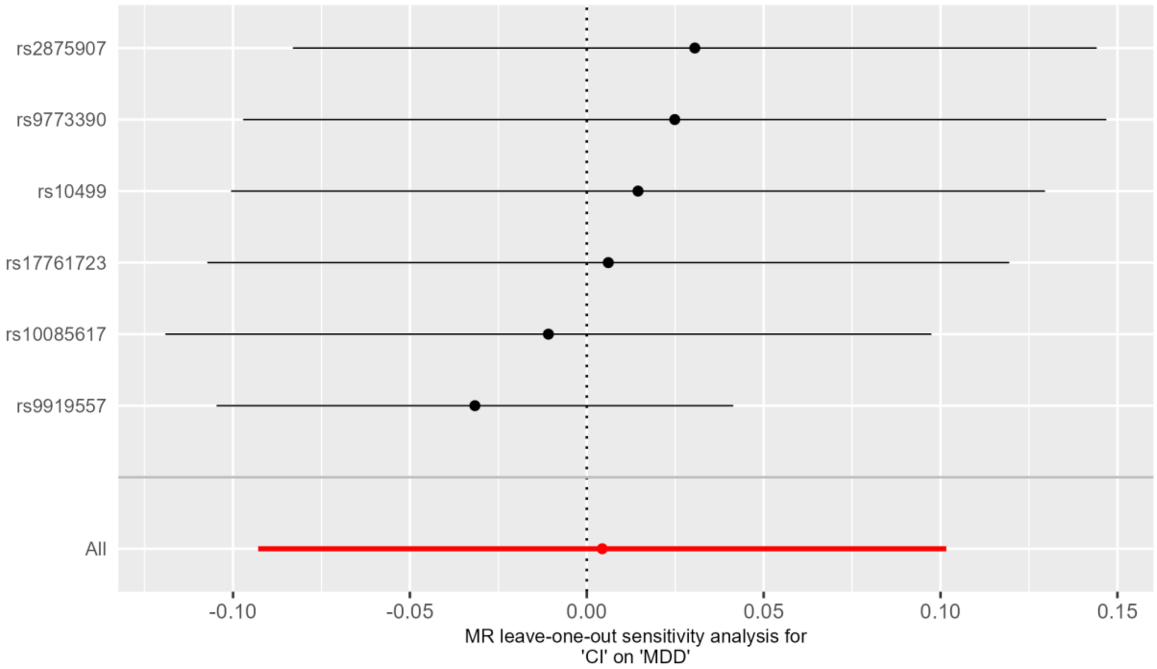

**Supplementary Figure S14.** Leave-one-out IVW regression analyses of liability to CUD on MDD risk.

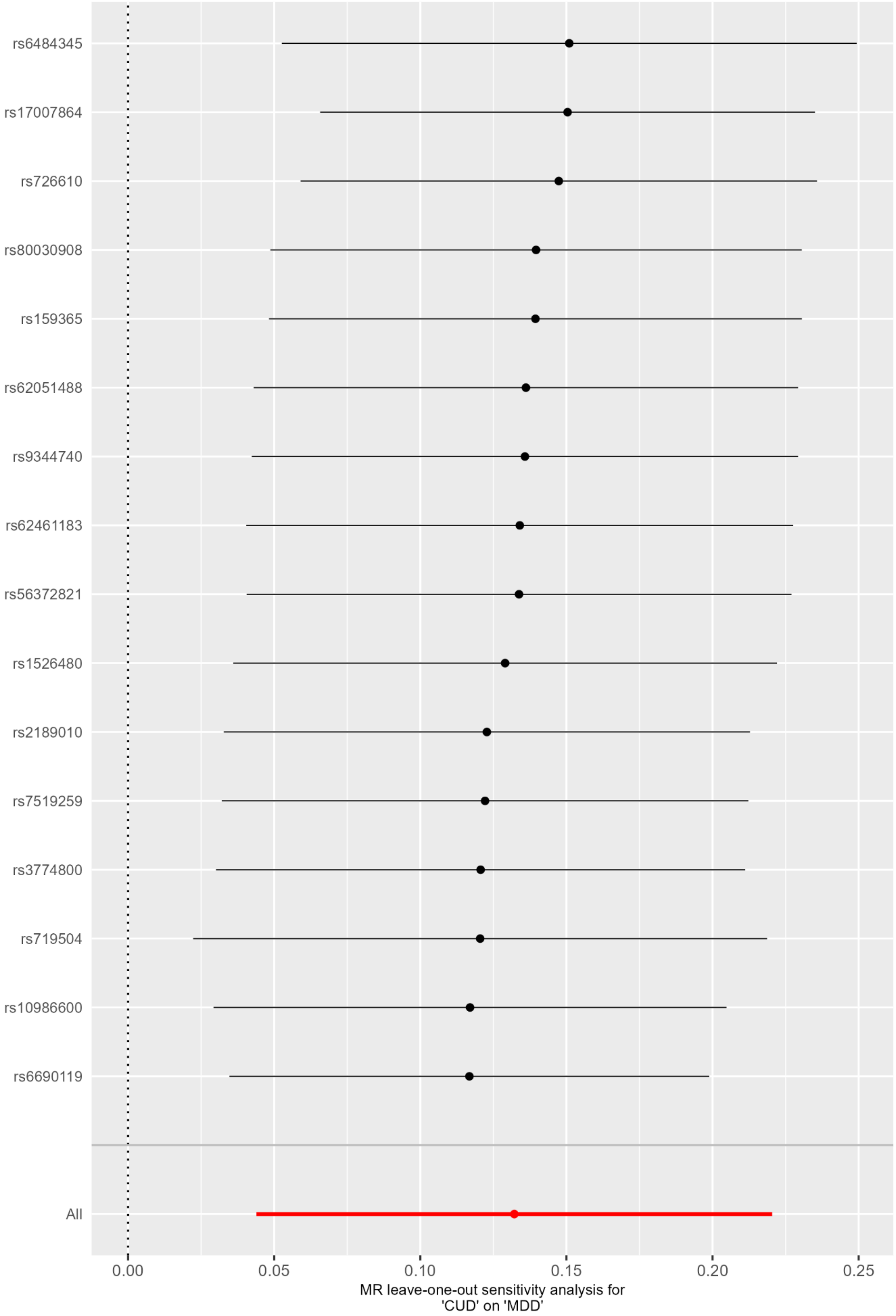

Supplementary Figure S15. Forest plot comparing univariable and MVMR effects of smoking initiation, cannabis initiation and CUD on MDD with Q-minimisation.

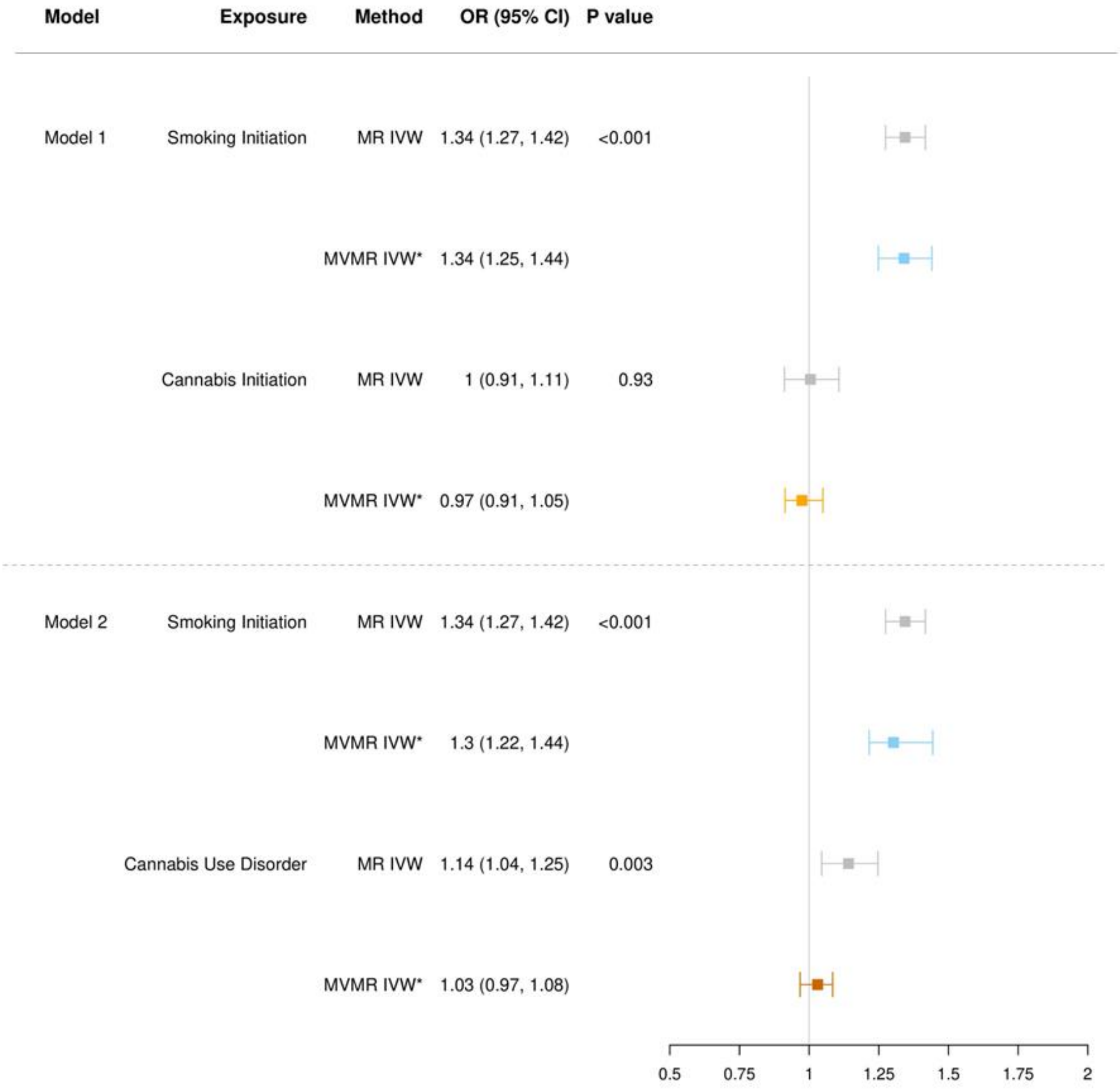

*Note.* **OR** = Odds Ratio; **CI** = Confidence Interval. ORs are scaled to per standard deviation increase in genetic liability to the exposure. Effect estimates are reported on the log odds scale with 95% confidence intervals. Robust MVMR estimates (MVMR IVW*) are depicted in colour, univariable estimates (MR IVW) are depicted in grey. **Model 1** refers to MVMR with smoking initiation and cannabis initiation as exposures. **Model 2** refers to MVMR with smoking initiation and CUD as exposures.

Supplementary Figure S16. Forest plot depicting univariable MR of the effect of smoking continuation and smoking heaviness on MDD in never smokers.

| Exposure | Method | OR (95% CI) | P value |
|---|---|---|---|
| Smoking Continuation | MR−IVW | 0.99 (0.64, 1.54) | 0.962 |
| | Weighted median | 1 (0.57, 1.73) | 0.989 |
| | Weighted mode | 0.98 (0.47, 2.05) | 0.955 |
| | MR−Egger (SIMEX) | 0.88 (0.53, 1.46) | 0.621 |
| | MR−PRESSO | 0.99 (0.82, 1.2) | 0.915 |
| Smoking Heaviness | MR−IVW | 1.11 (0.71, 1.72) | 0.654 |
| | Weighted median | 1.05 (0.55, 1.98) | 0.887 |
| | Weighted mode | 1.06 (0.56, 1.98) | 0.863 |
| | MR−Egger | 0.91 (0.42, 1.96) | 0.808 |
| | MR−PRESSO | 1.11 (0.94, 1.29) | 0.221 |

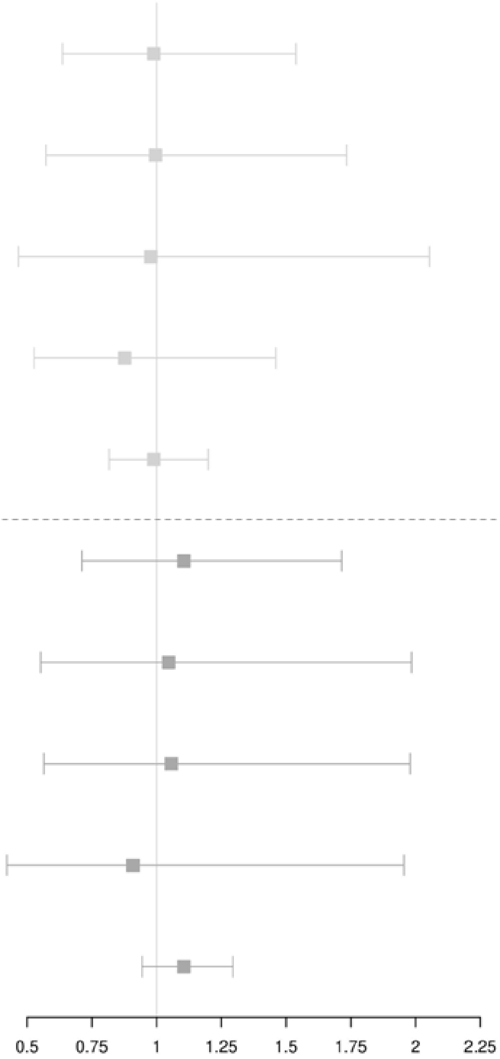

*Note.* **OR** = Odds Ratio; **CI** = Confidence Interval. ORs are scaled to per standard deviation increase in genetic liability to the exposure. Effect estimates are reported on the log odds scale with 95% confidence intervals. For MR-PRESSO, results present are 'raw estimates' where no outliers were identified, and 'outlier corrected' (OC) where outliers were identified. For MR-Egger, when $I^2_{GX}$ was 0.6-0.9, an unweighted SIMEX correction was applied, while estimates are not reported at all when $I^2$ was <0.6.
